# Selective reporting of outcomes and results in interrupted time series studies of health interventions: a methodological study

**DOI:** 10.1101/2025.11.06.25339702

**Authors:** Phi-Yen Nguyen, Simon L. Turner, Elizabeth Korevaar, Matthew J. Page, Joanne E. McKenzie

## Abstract

**Objective:** Selective reporting bias occurs when authors report outcomes or results based on the P value, magnitude or direction of the results. Selective reporting has not been examined in interrupted time series (ITS) studies. Therefore, we investigated (1) discrepancies in outcome reporting, (2) completeness of results reporting, and (3) evidence of selective reporting bias in ITS studies.

**Design:** We systematically searched for published peer-reviewed protocols of ITS studies of health interventions in 22 databases, and corresponding results reports addressing the protocol’s primary ITS research questions. For each primary research question, we identified outcomes that were reported in the protocol and the corresponding results report(s), and all results reported for those outcomes.

**Main outcome measure:** We defined a discrepancy as any outcome that was added, omitted, or had its primacy reclassified (e.g., from primary to secondary) in the results reports compared to the protocol. Each result was classified as fully reported if it was reported with both an effect estimate and a measure of precision (such as a confidence interval). Each result was also classified as favourable or not favourable to the interruption, based on its statistical significance and direction of effect.

**Results:** Our search for ITS protocols returned 4,590 abstracts. After excluding ongoing studies, protocols without published results for the primary research question, and records excluded for other reasons, we identified 44 ITS protocols (published 2010-2022) with 46 corresponding results reports. Among outcomes assessed for discrepancies, 52% (202/388) had a discrepancy, affecting 74% (31/42) of studies. Non-reporting of outcomes was prevalent, with 24% (132/553) of outcomes defined in the protocol not reported in the results report, and 60% (25/42) of protocols having at least one outcome not reported in the result reports. Only 28% (56/202) of discrepancies were justified by authors in the results reports. The association between a result favouring the interruption (based on statistical significance and direction of effect) and the result being fully reported was uncertain (OR=1.06 [95% CI 0.74 to 1.53]).

**Conclusion:** Non-reporting of outcomes and discrepancies in outcome reporting were prevalent. Pre-specifying outcomes in protocols and registries helps mitigate selective reporting. However, outcomes should be described in sufficient detail for readers to detect any changes.

**SUMMARY:** 

**Section 1: What is already known on this topic:** Selective outcome and result reporting bias ― occurs when the authors report a subset of the measured outcomes or results, based on whether the P value, magnitude of effect estimate or direction of effect supports the authors’ hypothesis. Selective reporting bias has not been examined in interrupted time series (ITS) studies.

**Section 2: What this study adds:** Our study highlights several issues with outcome reporting among ITS studies. We found that non-reporting of outcomes and discrepancies between outcomes reported in study protocols and outcomes reported in the final reports were prevalent. Moreover, outcomes were often too broadly defined in the protocol to allow an accurate assessment of selective reporting. We introduced a template for the elements that could be reported when describing an outcome in the context of ITS data.

**Section 3: How might these results change the focus of research or clinical practice?:** Our findings call for greater transparency in outcome reporting among investigators of ITS studies. To mitigate selective reporting bias, authors are encouraged to pre-specify outcomes in a protocol and describe outcomes in sufficient detail in any publication (protocol or results reports) to enable readers to detect any change in outcome reporting. Reporting may also be improved with guidance and stricter enforcement of outcome reporting by journal editors, peer reviewers and funders.

## INTRODUCTION

An interrupted time series (ITS) study is a non-randomised design commonly used to evaluate interventions that target populations for which randomisation may be unethical, infeasible or impractical. The design is a key method for evaluating “natural experiments” in which changes to infrastructure, polices or services are introduced by governments and healthcare systems (1).

In an ITS study, measurements of an outcome variable are often collected continuously over time and aggregated using summary statistics (e.g., means) within regular time intervals (e.g., monthly) for analysis. An ‘interruption’ separates the time series into pre- and post-interruption segments. Different models can be fitted to this data; most commonly, segmented regression models (2–4). Using this model, the underlying time trend in the aggregated outcome during the pre-interruption segment is estimated and extrapolated into the post-interruption segment, providing a counterfactual for what would have occurred in the absence of the interruption. This counterfactual is then compared with the trend estimated from the aggregated data in the post-interruption segment, from which various effect measures can be calculated; for example, immediate change in level at the time of the interruption and the difference in pre- and post-interruption slopes (5) (Figure 1) (5–7). The segmented regression model can be extended to adjust for additional features such as autocorrelation, seasonality, and time-varying confounders (7–9)..

**Figure 1.**
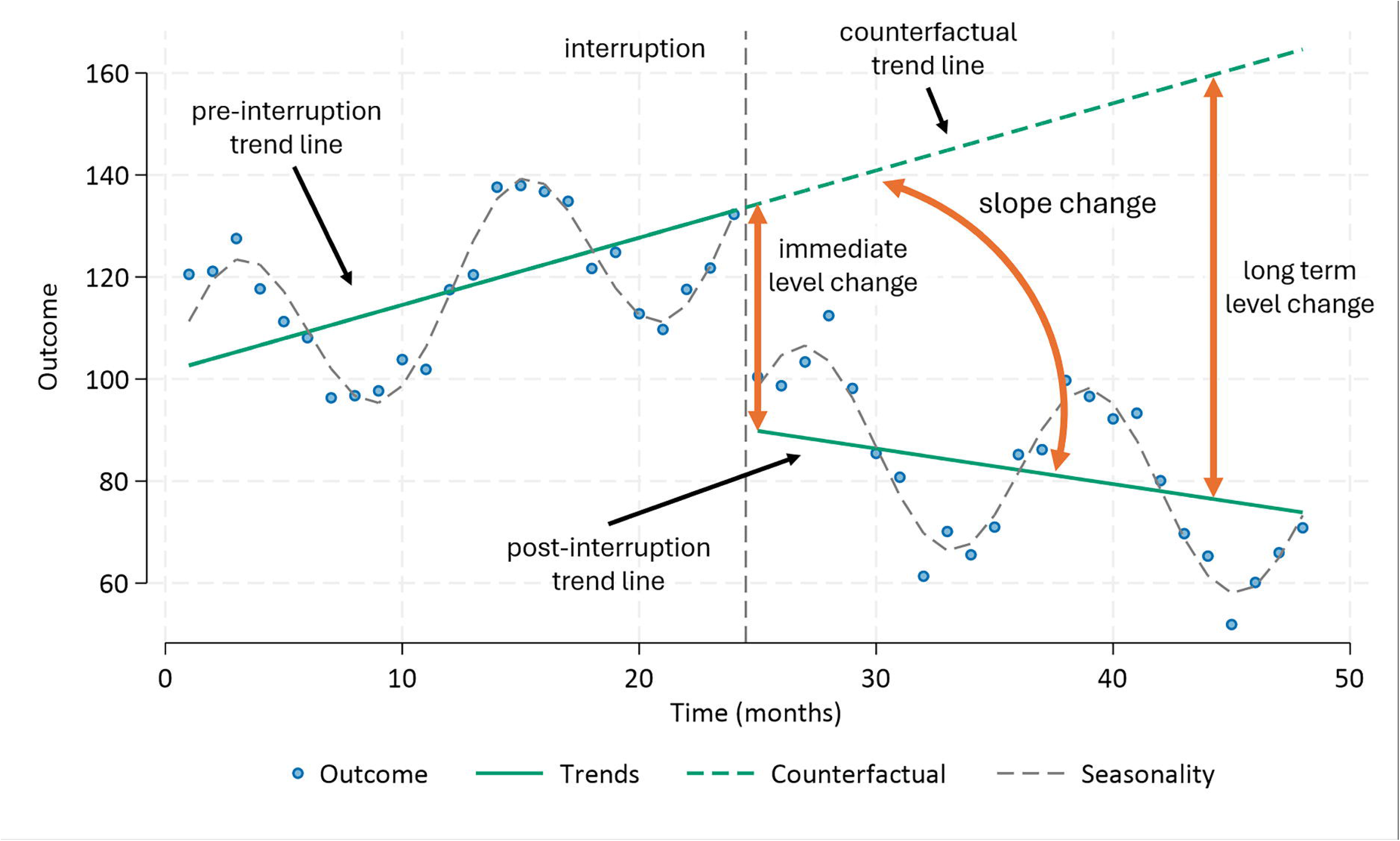
A graph of an interrupted time series.

The ITS design is less susceptible to selection and confounding biases, compared with other non-randomised designs (5). For this and other reasons, the design has seen an increase in popularity in recent times (3,4,10), with approximately 1000 studies published up to the year 2000, and 19,000 published since. In systematic reviews that examine the effects of interventions that target populations, ITS studies can provide valuable evidence, particularly when evidence from randomised trials is limited or unavailable (11).

Reporting bias can affect the validity of findings from systematic reviews. One common type of reporting bias ― selective outcome and result reporting bias ― occurs when the authors report a subset of the measured outcomes or analyses, based on whether the effect estimate’s magnitude, direction or P value supports the authors’ hypothesis (12–14). Withholding outcomes and results from publication can skew the evidence that informs systematic reviews and meta-analyses, potentially changing their conclusions about the benefits and harms of an intervention (15). Understanding the extent of selective reporting bias in ITS studies is important so that review authors, and users of ITS studies, are aware of this risk of bias when evaluating and interpreting evidence. Although selective reporting bias has been extensively investigated in samples of systematic reviews (16,17), randomised trials (18–20), and non-randomised studies (19,21), it has not been examined in ITS studies specifically.

## OBJECTIVES

- To examine the discrepancies between outcomes specified in ITS study protocols and those reported in the corresponding results reports;
- To examine how completely results are reported in ITS studies;
- To examine whether completeness of reporting is influenced by the statistical significance and direction of the results.

## METHODS

This study is one of three studies investigating reporting biases among ITS studies, for which we have published a protocol (22) (Supplementary File S1). Here, we provide an overview of the methods and the results for the second study; studies one and three are published separately. Deviations from the methods outlined in the protocol are available in Supplementary File S2, and the data extraction form is available in Supplementary File S3.

### 1. Creating a database of ITS studies

We searched for published peer-reviewed protocols of ITS studies from 22 databases, using a highly sensitive search filter for identifying ITS studies (23), as well as the corresponding results reports that addressed the primary ITS research question(s) from the protocol (see Supplementary Files S4 and S5 for details of search strategies, eligibility criteria, and processes for screening and determining the primary research question). ITS studies for which a protocol and a corresponding results report was found constituted the sample for this study.

### 2. Identifying, classifying and matching outcomes from protocols and results reports

#### 2.1. Identifying eligible outcomes

For each protocol and corresponding results report, two authors (PYN and one of SLT/EK/MJP) independently extracted all outcomes pertaining to the primary ITS research question, with outcomes extracted separately from the protocol and from the results report. We extracted: available details of the outcome (e.g., measurement tool, timing of outcome measurement, time interval of aggregation), and, whether the outcome was an impact outcome (i.e. expected to be affected by the interruption) or a control outcome (i.e. not expected to be affected by the interruption (24)). Unless otherwise specified, when we use the term outcome, we are referring to “impact outcomes”.

We adapted a previous framework to define outcomes in randomised control trials (RCTs) (25,26) so that it was more suitable for outcomes analysed in ITS studies. For each outcome, we summarised, when reported, the following five elements: domain, specific measure, time points, data type of individual measurements, and data type at aggregation (Table 1).

**Table 1.**
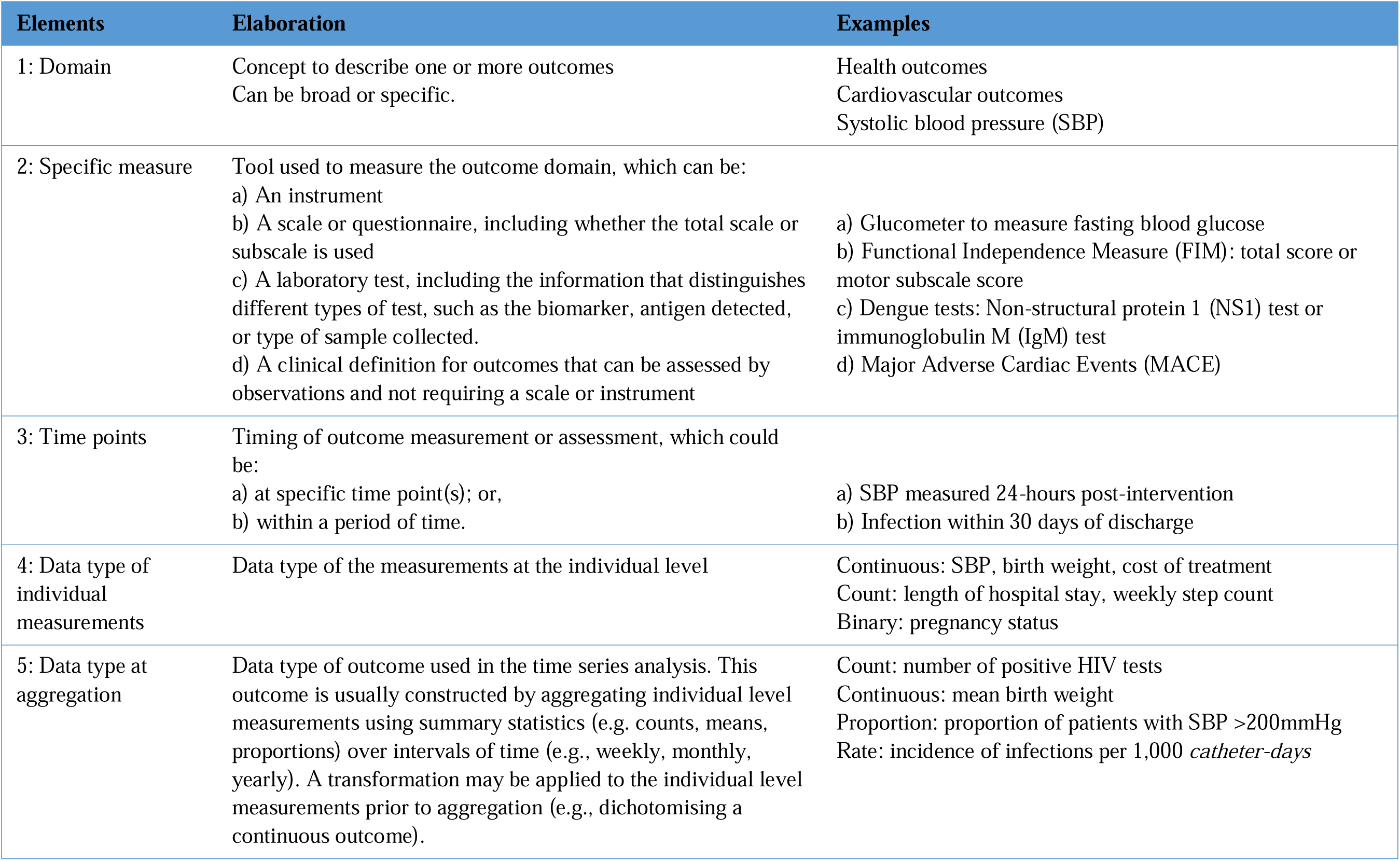
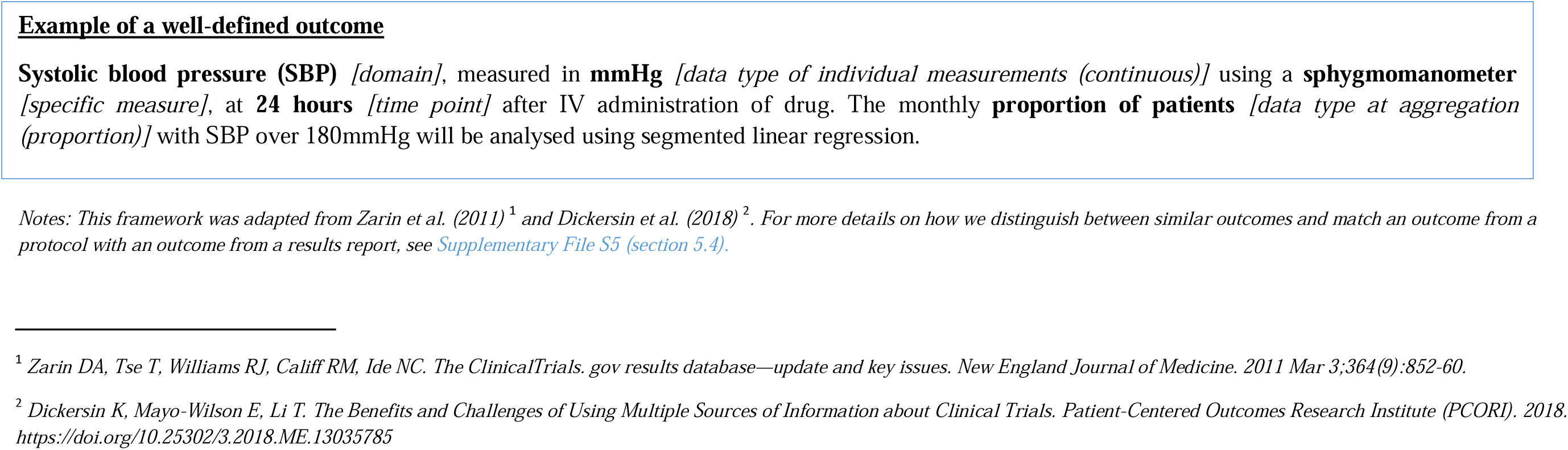
Framework for outcome definition.

We classified the primacy of each outcome as:

- *primary*, if the authors labelled it as such, or the outcome was used in a sample size calculation, or it was the only outcome in the study;
- *secondary,* if the authors labelled it as such;
- *could not be classified,* if the authors did not specify if the outcome was primary or secondary.

#### 2.2. Matching outcomes from protocol to report of result

One author (PYN) assessed whether each outcome from a protocol could be matched with a corresponding outcome from the results report, based on elements of outcome definition (see Section 2.1 and Supplementary File S5, section 5.4 for details). Only elements that were present in both documents were assessed – a match was confirmed if this subset of common elements were in agreement between the protocol and the results report. Each outcome in the protocol could be matched in the results report to a single outcome (*one-to-one*), to multiple outcomes (*one-to-many*, for example, ‘sale of opioids’ is specified in the protocol, with sales of the specific opioids ‘oxycodone’, ‘codeine’, and ‘fentanyl’ specified in the results report), or to *none at all*. Any complex cases of matching were discussed and resolved at team meetings (PYN/JEM/SLT/EK/MJP).

### 3. Identifying and extracting data on reported results

Two authors (PYN and one of JEM/SLT/EK/MJP) extracted all eligible results pertaining to the included outcomes from each results report. Discrepancies were resolved through discussion. An “eligible result” was defined as any measure of the difference between two segments of interest, including: a numerical result (e.g., an effect estimate such as an immediate level-change, with its confidence interval (CI), standard error (SE), or P value), or a qualitative statement about the statistical significance and/or the direction of the result (e.g., “There was no significant change in road fatalities after the policy was implemented”). For each eligible result, we extracted all numerical results reported, or if no numerical result was presented, any qualitative statement about statistical significance and/or the direction of effect. Note that multiple results per outcome were possible arising from different effect measures (e.g., immediate level change, slope change).

Using the approach proposed by Chan and colleagues (4), we classified each result as:

- *fully reported*, if sufficient data were reported to include a result in a meta-analysis, that is, an effect estimate and a measure of precision (CI or SE);
- *partially reported*, if insufficient data were reported to include a result in a meta-analysis (e.g., an effect estimate is reported without any measure of precision); or
- *qualitatively reported,* if only a qualitative statement or a P value was reported.

Lastly, where possible, we classified each result as being favourable to the interruption or not. A result was classified as “favourable” if both (i) the direction of the effect was favourable to the interruption (i.e., the effect estimate indicated greater benefit or less harm compared with the counterfactual), and (ii) the result was statistically significant (i.e. P value <0.05, or if absent, the 95% CI excluded the null, or the authors stated the result was statistically significant). Results that did not meet either criteria were classified as “not favourable”.

### 4. Assessing discrepancies in outcome reporting

Control outcomes and outcomes with one-to-many matches were excluded from the assessment of discrepancies. We excluded outcomes with one-to-many matches because it was impossible to establish from the protocol whether the matched outcomes were ever intended to be included.

We defined a discrepancy in outcome reporting as any of the following (full details in Supplementary File S5, section 5.4):

- *missing in results report*, where an outcome specified in the protocol was not mentioned in the report of the results;
- *missing in protocol*, where an outcome mentioned in the report of the results was not pre-specified in the protocol;
- *discrepancy in outcome primacy*, where a primary outcome in the protocol was demoted to secondary or unclassified outcome in the results report; or, a secondary or unclassified outcome in the protocol was promoted to primary in the results report.

### 5. Statistical analysis

We summarised results by calculating frequencies and percentages for categorical variables (e.g., results that are fully reported, partially reported, qualitatively reported) and medians and interquartile ranges (IQR) for continuous variables. In addition, we summarised and categorised reported reasons for outcome discrepancies. We conducted a sensitivity analysis to examine the impact on the percentage of outcomes with a discrepancy by assuming that all outcomes for which authors provided a justification were not discrepancies. This provides a “best-case scenario” for the magnitude of outcome discrepancy, as it assumes that all the justifications are valid, which may not be the case.

To examine whether there was an association between a result being favourable to the interruption and its completeness of reporting (fully reported / partially or qualitatively reported), we fitted a marginal logistic regression model using generalised estimating equations, with an exchangeable working correlation, robust standard errors clustered by study, and fixed effects for: whether a result is favourable to the interruption or not, and potential confounders: type of funding (industry, non-industry [government, education institutions or not-for-profit entities] or no funding [as declared by authors]) (27) and outcome primacy (primary or secondary / unclassified) (16). Results are reported as odds ratios (OR) with 95% CIs. Analyses were undertaken in Stata version 18 (28).

## RESULTS

### 1. Overview of the search process & extracting outcomes/results

Of the 148 full texts retrieved from the literature search, 120 met the inclusion criteria for an eligible ITS study protocol (Figure 2). A search for results reports matching these 120 protocols identified 467 potentially eligible full texts, from which 46 met our inclusion criteria as eligible results reports. Of the 120 ITS studies with protocols, 44 studies (corresponding to 44 protocols and 46 results reports) constituted the sample for this study. From these studies, we identified 572 eligible outcomes and 860 eligible results corresponding to the identified primary ITS research question (Figure 2).

**Figure 2.**
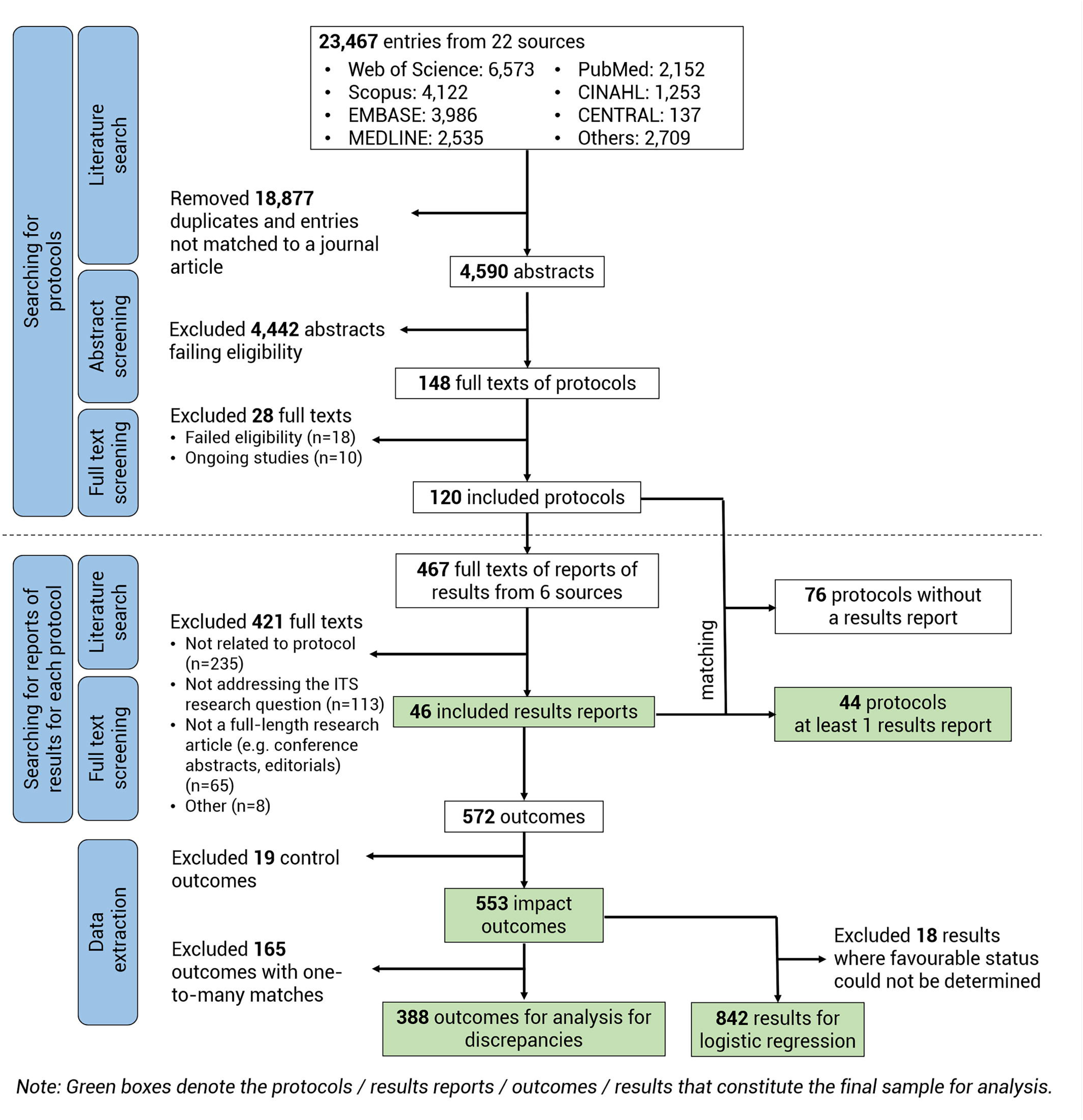
Flowchart showing processes of literature search and screening.

### 2. Characteristic of included studies

Most studies were supported by non-industry funding (37/44, 84%), most were conducted in high-income countries (36/44, 82%), and the majority examined practice change interventions in a clinical setting (25/44, 57%) (Table 2). Half of the studies were registered (22/44, 50%). Three-quarters of the studies outlined methods for other study designs in addition to the ITS study analysis (e.g., process evaluations) (33/44, 75%).

**Table 2.** Characteristics of included studies.

| Characteristic | No. ITS studies (%)<br>(N=44) |
| --- | --- |
| Focus of the study |  |
| Only ITS study | 11 (25%) |
| ITS and other study designs | 33 (75%) |
| Type of funding |  |
| No funding | 1 (2%) |
| Non-industry funding | 37 (84%) |
| Industry funding | 6 (14%) |
| Study registration | 22 (50%) |
| Nature of the interruption |  |
| Exposure (natural events) | 0 (0%) |
| Intervention | 44 (100%) |
| Practice change in a clinical setting | 25 (57%) |
| Health system interventions | 8 (18%) |
| Policy & regulatory changes | 6 (14%) |
| Social & economic interventions | 3 (7%) |
| Environmental interventions | 2 (5%) |
| Other | 0 (0%) |
| Level at which the intervention was implemented |  |
| Unit-based or institutional | 18 (41%) |
| Regional | 8 (18%) |
| National | 17 (39%) |
| Multinational | 1 (2%) |
| Level at which the intervention was evaluated |  |
| Unit-based or institutional | 21 (48%) |
| Regional | 10 (23%) |
| National | 12 (27%) |
| Multinational | 1 (2%) |
| Country where study was conducted<sup>†</sup> |  |
| High-income countries | 36 (82%) |
| Upper middle-income countries | 3 (7%) |
| Lower middle-income countries | 4 (9%) |
| Low-income countries | 1 (2%) |
| Timing of data collection relative to the protocol's submission |  |
| Retrospective | 17 (39%) |
| Prospective | 27 (61%) |

Notes: For details of how we defined the options for each characteristic, refer to Supplementary File S3 (Data extraction form). Abbreviation: ITS: interrupted time series
<sup>†</sup>Based on World Bank Group's FY25 income classification. Total of percentages may exceed 100% as multiple response options could apply.

### 3. Characteristics of included outcomes

We extracted 572 outcomes from the included studies; 516/572 (90%) outcomes were specified in the protocols and 433/572 (76%) outcomes were specified in the results report. Nearly all outcomes were “impact” outcomes (553/572, 97%), while 19/572 (3%) were “control” outcomes. 234/572 (41%) outcomes were one-to-one matched (i.e., an outcome in the protocol matched with a single outcome from the results report) and 165/572 (29%) were one-to-many matched (i.e., an outcome in the protocol matched with multiple outcomes in the results report). The remaining 173/572 (30%) had no match, including 117/572 (20%) outcomes that appeared only in the protocol and 56/572 (10%) outcomes that appeared only in the results report (Table 3).

**Table 3.**
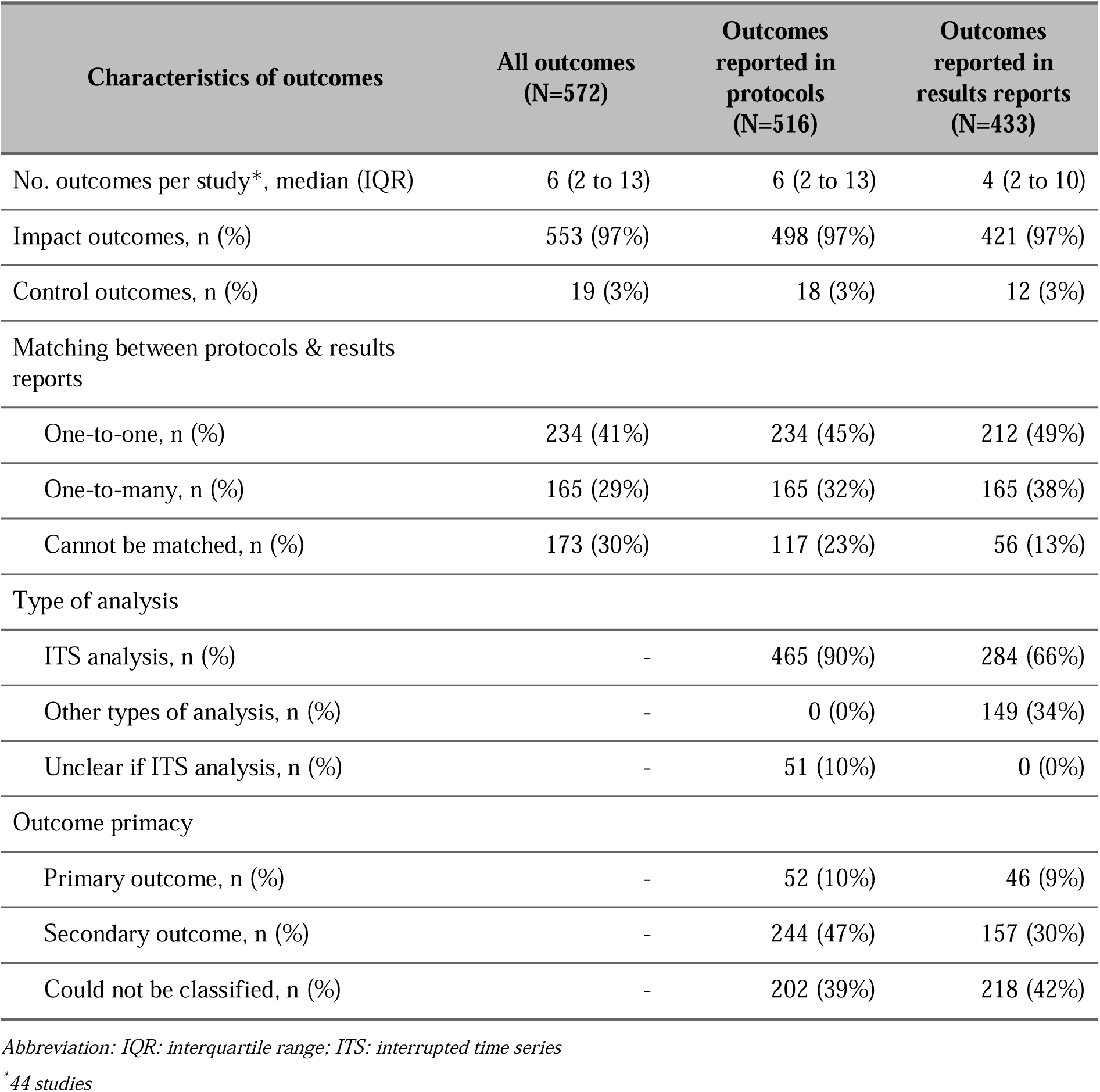
Characteristics of included outcomes.

| Characteristics of outcomes | All outcomes<br>(N=572) | Outcomes<br>reported in<br>protocols<br>(N=516) | Outcomes<br>reported in<br>results reports<br>(N=433) |
| --- | --- | --- | --- |
| No. outcomes per study*, median (IQR) | 6 (2 to 13) | 6 (2 to 13) | 4 (2 to 10) |
| Impact outcomes, n (%) | 553 (97%) | 498 (97%) | 421 (97%) |
| Control outcomes, n (%) | 19 (3%) | 18 (3%) | 12 (3%) |
| Matching between protocols & results reports |  |  |  |
| One-to-one, n (%) | 234 (41%) | 234 (45%) | 212 (49%) |
| One-to-many, n (%) | 165 (29%) | 165 (32%) | 165 (38%) |
| Cannot be matched, n (%) | 173 (30%) | 117 (23%) | 56 (13%) |
| Type of analysis |  |  |  |
| ITS analysis, n (%) | - | 465 (90%) | 284 (66%) |
| Other types of analysis, n (%) | - | 0 (0%) | 149 (34%) |
| Unclear if ITS analysis, n (%) | - | 51 (10%) | 0 (0%) |
| Outcome primacy |  |  |  |
| Primary outcome, n (%) | - | 52 (10%) | 46 (9%) |
| Secondary outcome, n (%) | - | 244 (47%) | 157 (30%) |
| Could not be classified, n (%) | - | 202 (39%) | 218 (42%) |

Abbreviation: IQR: interquartile range; ITS: interrupted time series
\* 44 studies

### 4. Characteristics of included results

We identified 860 eligible results corresponding to the identified primary ITS research question, with a median of 11 results (IQR 4 to 26) per study and 1 result (IQR 0 to 3) per outcome. Of the 860 results, 201/860 results (23%) were classified as favourable to the interruption, 641/860 results (75%) were classified as not favourable to the interruption, and 18/860 (2%) could not be classified due to lack of information. The median percentage of results favourable to the interruption per study was 20% (IQR 3% to 50%) (Table 4).

**Table 4.** Patterns and discrepancies in reporting of outcomes.

|  | No. outcomes (%) | No. outcomes per study, median (IQR) (44 studies) | Percentage of outcomes per study, median (IQR) (44 studies) |
| --- | --- | --- | --- |
| **By completeness of reporting at the outcome level<sup>†</sup> (n=553)** |  |  |  |
| All results were fully reported | 315 (57%) | 1 (0 to 4) | 27% (0% to 100%) |
| At least one result was fully reported but not all | 47 (8%) | 0 (0 to 0) | 0% (0% to 0%) |
| At least one result was partially reported, but none was fully reported | 12 (2%) | 0 (0 to 0) | 0% (0% to 0%) |
| Only qualitatively reported results | 47 (8%) | 0 (0 to 0) | 0% (0% to 0%) |
| No result was reported | 132 (24%) | 1 (0 to 4) | 16% (0% to 46%) |
| **By type of discrepancies (n=388\*)** |  |  |  |
| No discrepancy | 186 (48%) | 2 (0 to 5) | 40% (0% to 83%) |
| Discrepancy | 202 (52%) | 2 (0 to 6) | 27% (0% to 71%) |
| Missing in protocol | 55 (14%) | 0 (0 to 0) | 0% (0% to 3%) |
| Missing in results report | 132 (34%) | 1 (0 to 4) | 14% (0% to 46%) |
| Discrepancy in outcome primacy | 15 (4%) | 0 (0 to 0) | 0% (0% to 0%) |
| Primary outcome was demoted to secondary/unclassified outcome | 4 (1%) | 0 (0 to 0) | 0% (0% to 0%) |
| Secondary/unclassified outcome was promoted to primary outcome | 11 (3%) | 0 (0 to 0) | 0% (0% to 0%) |

Abbreviation: IQR: interquartile range; ITS: interrupted time series
<sup>†</sup>Each outcome can have multiple results. For example, after fitting a segmented linear regression, the authors can report two results for each outcome: an estimate of the slope change and an estimate of the immediate level change.
\*Excluding outcomes with one-to-many matches and control outcomes.

### 5. Completeness in reporting at the study, outcome and result level

At the study level, in 12/44 (27%) studies, all of the outcomes had all of their results fully reported; in 7/44 (16%) studies, all of the outcomes had reported results, but to a varying degree of completeness. The remaining 25/44 (57%) studies had at least one outcome without any reported result (Figure 3).

**Figure 3.**
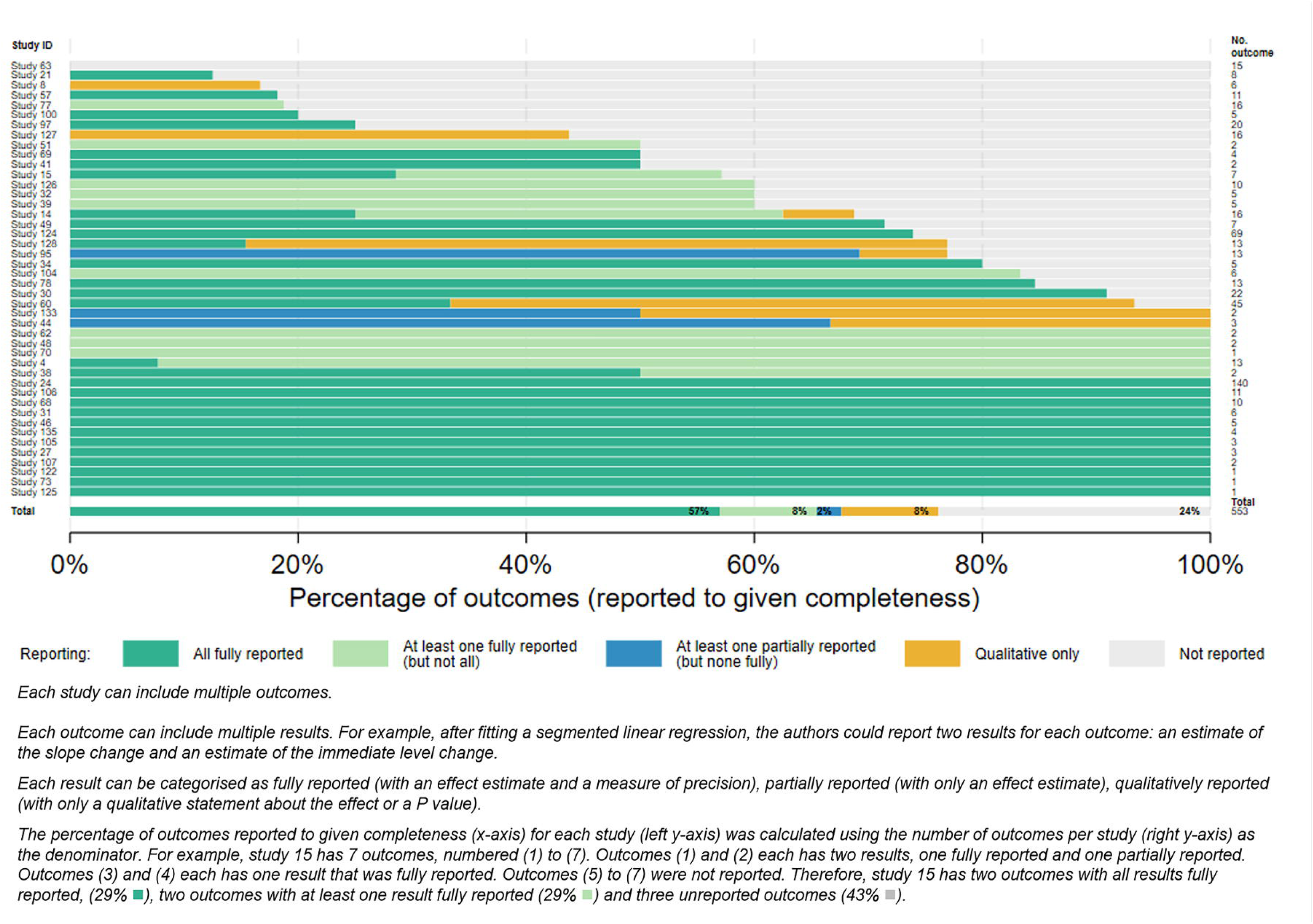
Bar plot showing proportion of outcomes reported to different levels of completeness per study and overall.

At the outcome level, more than half of the outcomes had all of their results fully reported (315/553, 57%); 47/553 (8%) had at least one fully reported result but not all; 12/553 (2%) had only partially reported results but none that were fully reported, and 47/553 (8%) had only qualitatively reported results. A quarter of the outcomes (132/553, 24%) had no eligible results reported, or were not mentioned in the results report at all (Table 4).

At the result level, most of the results presented in the results reports were fully reported (741/860, 86%), with a median of 6 fully reported results per study (IQR 4 to 18) and 1 fully reported result per outcome (IQR 0 to 2). A small number of results were partially reported (28/860, 3%), and 91/860 results (11%) were qualitatively reported (Table 5). In 31/44 (70%) studies, all of the reported results were fully reported; in the remaining 10/44 (23%) studies, the reported results were reported to varying degree of completeness (Figure 4). Two studies only had qualitatively reported results and one study had no eligible result.

**Figure 4.**
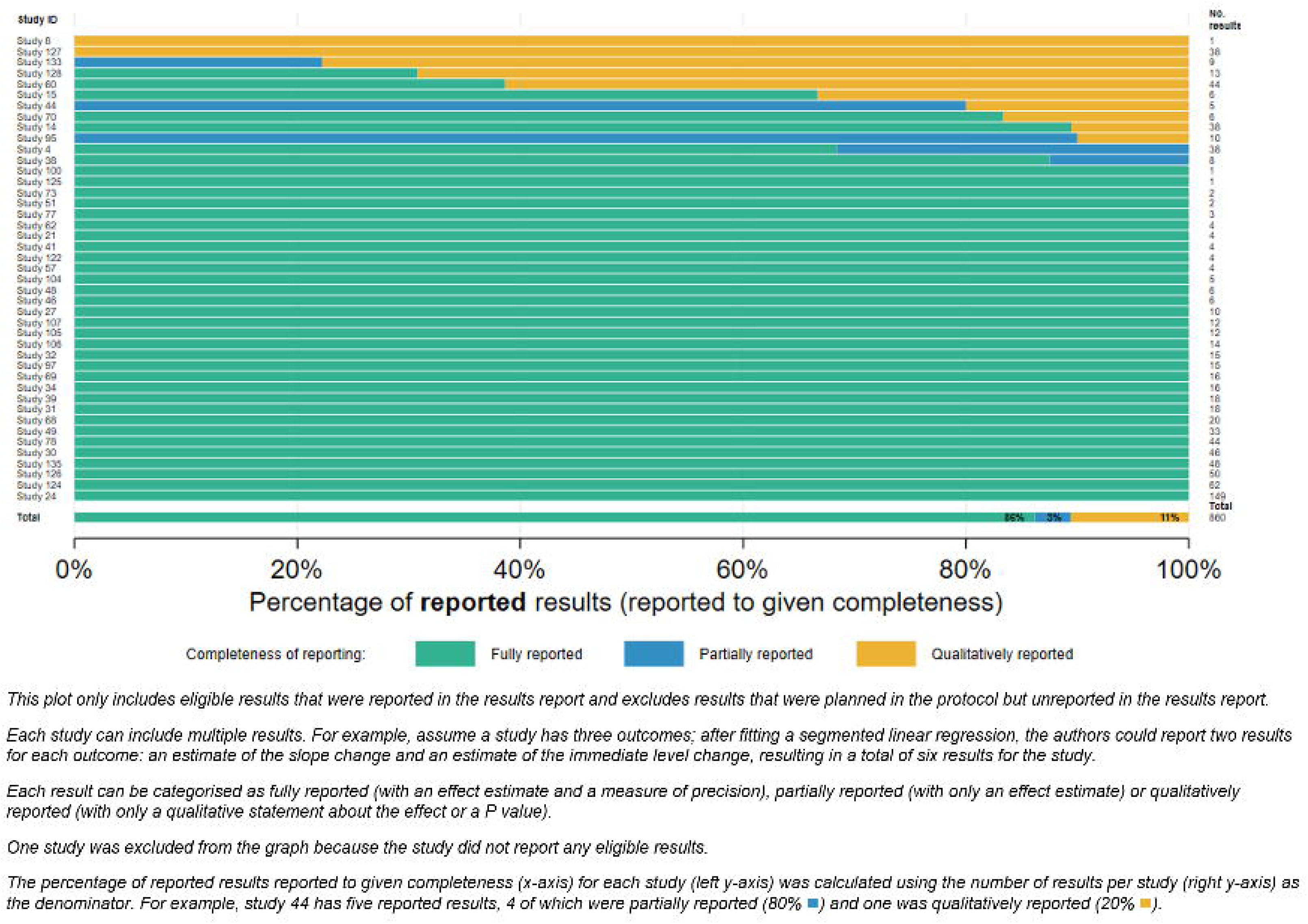
Bar plot showing percentage of results reported to different levels of completeness per study and overall.

**Table 5.** Characteristics of results.

|  | No. results (%)<br>(N=860) | No. results per results<br>report, median (IQR)<br>(44 studies) | Percentage of results per<br>results report, median (IQR)<br>(44 studies) | No. results per outcome,<br>median (IQR)<br>(572 outcomes) | Percentage of results per<br>outcome, median (IQR)<br>(572 outcomes) |
| --- | --- | --- | --- | --- | --- |
| Completeness of reporting |  |  |  |  |  |
| Fully reported | 741 (86%) | 6 (4 to 18) | 100% (85% to 100%) | 1 (0 to 2) | 100% (0% to 100%) |
| Partially reported | 28 (3%) | 0 (0 to 0) | 0% (0% to 0%) | 0 (0 to 0) | 0% (0% to 0%) |
| Qualitatively reported | 91 (11%) | 0 (0 to 0) | 0% (0% to 0%) | 0 (0 to 0) | 0% (0% to 0%) |
| Statistical significance of the<br>result |  |  |  |  |  |
| Significant | 283 (33%) | 4 (1 to 10) | 32% (21% to 61%) | 0 (0 to 1) | 0% (0% to 38%) |
| Not significant | 536 (62%) | 6 (2 to 12) | 56% (33% to 75%) | 1 (0 to 1) | 50% (0% to 100%) |
| Cannot be determined | 41 (5%) | 0 (0 to 0) | 0% (0% to 0%) | 0 (0 to 0) | 0% (0% to 0%) |
| Direction of the result |  |  |  |  |  |
| Favourable to the interruption | 471 (55%) | 6 (2 to 12) | 58% (41% to 83%) | 0 (0 to 1) | 0% (0% to 100%) |
| Favourable to the<br>comparator/neither side | 348 (40%) | 4 (1 to 8) | 33% (7% to 55%) | 0 (0 to 1) | 0% (0% to 100%) |
| Cannot be determined | 41 (5%) | 0 (0 to 0) | 0% (0% to 0%) | 0 (0 to 0) | 0% (0% to 0%) |
| Overall favourable status of the<br>result<sup>†</sup> |  |  |  |  |  |
| Favourable to the interruption | 201 (23%) | 2 (1 to 8) | 20% (3% to 50%) | 0 (0 to 0) | 0% (0% to 0%) |
| Not favourable to the<br>interruption | 641 (75%) | 8 (2 to 16) | 77% (43% to 91%) | 1 (0 to 1) | 96% (0% to 100%) |
| Cannot be determined | 18 (2%) | 0 (0 to 0) | 0% (0% to 0%) | 0 (0 to 0) | 0% (0% to 0%) |

Abbreviation: IQR: interquartile range
<sup>†</sup>The overall favourable status was determined based on two conditions: (A) the result was statistically significant (i.e. $P$ value $<0.05$ , or if absent, the 95% CI excluded the null, or the authors stated the result was statistically significant), and (B) the direction of the effect was favourable to the interruption (i.e. the effect estimate indicated greater benefit or less harm compared with the control).
- If both (A) and (B) are true, the result was classified as “favourable to the interruption”. - If either (A) and (B) is false, the result was classified as “not favourable to the interruption”. - If there is insufficient information to classify, such as when (A) is true and (B) is unclear, (B) is true and (A) is unclear, or both (A) and (B) are unclear, the result was classified as “could not be determined”.

### 6. Discrepancies in outcome reporting at the study and outcome level

After excluding 19 control outcomes and 165 outcomes with one-to-many matches, 388 outcomes from 42 studies were assessed for discrepancies. At the study level, 31/42 (74%) had at least one outcome with discrepancy and 11/42 (26%) studies had no discrepancy. More than half of the studies were affected by outcomes missing in the results report (25/42, 60%), followed by outcomes missing in the protocol (11/42, 26%), or by discrepancies in outcome primacy (10/42, 24%).

At the outcome level, approximately half (202/388, 52%) of the outcomes had a discrepancy in reporting. Specifically, 132/388 (34%) outcomes were specified in the protocol but missing in the results report; 55/388 (14%) outcomes were reported in the results report but missing in the protocol; 15/388 (4%) outcomes had a discrepancy in their outcome primacy. The remaining 186/388 outcomes (48%) were not discrepant. The median percentage of outcomes with any discrepancy per study was 27% (IQR 0% to 71%). The median percentage of outcomes per study with no discrepancy was 40% (IQR 0% to 83%) (Table 4).

Of the 202 outcomes with discrepancies, a justification was provided for the discrepancy for 56/202 (28%) outcomes from 11/31 (35%) studies (Table 6). No justification was provided for a change in primacy of the outcome for any outcome with this type of discrepancy. The most common reasons for not reporting an outcome specified in the protocol were that rates of the outcomes were too low to undertake the analysis (affecting 22/202 [11%] outcomes in 4 [13%] studies) and the outcome was deemed unsuitable in evaluating the effect of intervention (affecting 20/202 [10%] outcomes in 3 [10%] studies). Outcomes were added in the results reports for various reasons, primarily because data for originally planned outcomes could not be collected (affecting 5/202 [2%] outcomes in 1 [3%] study). Our sensitivity analysis, in which we considered justified outcome discrepancies to no longer be discrepancies, yielded 38% (146/388) of the outcomes with a discrepancy in reporting, affecting 69% (29/42) studies (Supplementary File S6).

**Table 6.** Justification for outcome discrepancies.

| **Reason** | **No. of outcomes affected (%)**<br>**(N=202<sup>a</sup>)** | **No. of studies affected (%)**<br>**(N=31<sup>b</sup>)** |
| --- | --- | --- |
| **For not reporting a planned outcome in the results report** |  |  |
| Rates of outcomes were too low to undertake the analysis | 22 (11%) | 4 (13%) |
| Outcome was deemed unsuitable in evaluating the effect of intervention | 20 (10%) | 3 (10%) |
| Data were missing (no specific reason given) | 3 (1%) | 2 (6%) |
| Outcome was not accurately measured | 2 (<1%) | 2 (6%) |
| Outcome was deemed unnecessary by authors | 2 (<1%) | 2 (6%) |
| **For adding a new outcome in the results report** |  |  |
| Outcome was added after data for originally planned outcomes could not be collected | 5 (2%) | 1 (3%) |
| Outcome was added after the original outcome was not accurately measured | 1 (<1%) | 1 (3%) |
| Outcome was added after the original outcomes were deemed unsuitable | 1 (<1%) | 1 (3%) |
| **Total** | **56 (28%)** | **11 (35%)<sup>c</sup>** |

<sup>a</sup>Total number of outcomes with discrepancies
<sup>b</sup>Total number of studies with discrepancies
<sup>c</sup>Total of number of studies may exceed 11 because some studies had more than one type of justification provided.

### 7. Selective reporting of results

Of the 201 results favourable to the interruption, 183 were fully reported (91%), while of the 641 results not favourable to the interruption, 558 were fully reported (87%). The adjusted odds ratio (estimated from the logistic regression model) was OR=1.06 (95% CI 0.74 to 1.53) (Supplementary File S7) suggesting that the odds of a result being fully reported was slightly larger for results favourable to researchers’ hypotheses, but that there was uncertainty in this association.

## DISCUSSION

### 1. Principal findings and comparison with the literature

Our study provides important insights about the extent of selective outcome reporting bias in ITS studies. We identified 44 ITS protocols and their published results, identifying 572 outcomes, and 860 results from these studies. Our findings highlight several issues with outcome reporting among ITS studies.

First, we observed a high frequency of discrepancies in outcome reporting between the study protocols and their corresponding final reports. Three quarters (74%) of the studies had at least one outcome with a discrepancy. More than half (52%) of the outcomes assessed had a discrepancy, higher than that observed in studies investigating outcome discrepancies among systematic reviews (38% (17)) and similar to that among randomised trials (54% (20)). The most prevalent type of discrepancy was the omission of outcomes that were planned in the protocol, accounting for 34% of the assessed outcomes. This type of discrepancy could importantly affect an evidence base if the decision not to report the planned outcome is made based on the nature of the results. Justification for changes in outcome reporting was infrequent (only 28% of outcomes with discrepancies were given justifications). Expectedly however, most of the justifications related to practical issues (e.g., issues with the data collection or implementation). Ascertaining other reasons for changes (e.g., whether it was based on statistical significance of the outcome, whether the researchers considered the findings relevant to research question or important for readers) (29), may be more appropriately investigated by surveying the ITS authors.

Second, many outcomes were too broadly defined in the protocol to allow assessment of discrepancies. Nearly 30% of outcomes in the protocol could be matched to multiple outcomes in the results reports; for example, in one protocol, the primary outcome was “the number of antibiotics prescriptions”, while the results report included separate outcomes for the number of prescriptions of different antibiotics (e.g., amoxicillin, cephalexin and ciprofloxacin). A broadly-defined outcome in the protocol leaves the door open for selective reporting, or presenting outcomes with significant findings as if they were planned outcomes (30). An example of a well-defined outcome, with reporting of each element, is provided in Table 1.

Third, non-reporting of outcomes is prevalent, with 57% of studies having at least one unreported outcome, and 24% of the outcomes having no results reported at all. The percentage of studies with unreported outcomes (57%) is slightly lower than that found among outcomes in randomised trials (60-88%) (31,32). We note that for these investigations, data from unreported outcomes was solicited from trial investigators (31,32), which was not done in our study. However, the frequency of incomplete reporting of outcomes in our study (18%) was substantially less compared with what has been found in randomised trials (31-65%).

Lastly, our examination of the nature of the results (i.e. the direction and statistical significance) on completeness of reporting was uncertain due to the width of the confidence interval. Studies examining this association in randomised trials (using a similar classification of reporting completeness) (31,32), found large associations between statistical significance and complete reporting of results (odds ratios ranging from 2.4 to 4.7). Moreover, compared to randomised trials, ITS studies are not required to be registered and there were fewer ITS protocols published. Researchers publishing protocols are perhaps more likely to be aware of, and thus more adherent to, reporting requirements, including completely reporting all results.

### 2. Implications for practice

Our findings suggest that authors of ITS studies do not report at least some of the planned outcomes. At best, this means that not all evidence is available for evidence synthesis; at worst, that the available evidence is biased, as will occur when the authors’ decision to report results has been based on the nature of those results. This highlights the need for registration of ITS studies and analyses, and publication of protocols and statistical analysis plans, which describe all outcomes in detail. Those who synthesise evidence from ITS studies should seek out registration details, a protocol or statistical analysis plan for each study, to assess for non-reporting of outcomes, even although they are not commonly reported (14). In addition, readers should be alert to missing outcomes that would be expected to be measured given the interruption and the context, such as blood pressures for interventions framed as “directed at individuals with hypertension” (14). Reporting sufficiently detailed outcomes will allow readers to detect any changes in the outcomes reported in the final publication. Our framework for defining an outcome in the context of ITS data provides a template for the elements that could be reported (Table 1).

It is inevitable that in some ITS studies, changes to outcomes will be required due to, for example, logistical or analytical challenges (e.g., planned data collection was not feasible). Reporting such changes, along with the rationale, is a recommendation in the CONSORT 2025 statement (item 10) (33), a reporting guideline for randomised trials that similarly applies to ITS studies. Stricter monitoring and enforcement of outcome reporting by journal editors, peer reviewers and funders may improve reporting of all planned outcomes (34).

### 3. Strengths and limitations

To our knowledge, this is the first study to investigate selective outcome reporting in ITS studies. Many studies have investigated outcome reporting bias in clinical trials (12,35,36) or systematic reviews of RCTs (17), but few have been undertaken for non-randomised studies, potentially due to the difficulty in creating an inception cohort (i.e. a set of studies known to have been initiated, irrespective of their results) (19). Our framework to define outcomes was based on a previous framework developed for outcomes in clinical trials (25,33). A key difference with ITS outcomes is that the individual measurements are typically aggregated (using summary statistics) over units of time (e.g., months). We introduced new elements of outcome definitions to accommodate this difference, which allowed us to match the outcomes more accurately and consistently between protocols and results reports.

Our study is not without limitations. Our sample includes only ITS studies with published protocols, which is a very small fraction of all ITS studies published. ITS studies with published protocols may be more completely reported than those without protocols. If this is true, our study might have underestimated the extent of outcome reporting discrepancies and incomplete reporting of results. Second, in many protocols, authors did not explicitly report what effect measures they planned to report. Therefore, we could not assess the extent of selective reporting at the result level (i.e. how many results were planned but not reported).

## CONCLUSION

Even among ITS studies with published protocols, which likely represent best practice, non-reporting of outcomes and discrepancies in outcome reporting were prevalent. Outcomes should be pre-specified in a protocol or registry to mitigate selective reporting, and described in sufficient detail in any publication (protocol or results reports) to enable readers detect any change in outcome reporting.

## Data Availability

All datasets and analytical code can be found on the Open Science Framework (DOI: osf.io/9jqh3 and osf.io/b837n).

https://osf.io/b837n

## ACKNOWLEDGEMENT

Ethics approval and informed consent

This study was conducted using publicly available data and did not involve human participants, identifiable personal data, animal subjects, or interventions requiring ethical approval. Data related to the included studies was deidentified and anonymised using unique identification numbers.

## Availability of data and materials

All datasets and analytical code can be found on the Open Science Framework (DOI: <u>osf.io/9jqh3 and</u> <u>osf.io/b837n</u>).

## Transparency statement

The authors declare that the manuscript is an honest, accurate, and transparent account of the study being reported; that no important aspects of the study have been omitted; and that any discrepancies from the study as originally planned have been reported in the Supplementary Files.

## Patient and Public Involvement

Patients and the public were not involved in the design and conduct of this methodological research study.

## Contributorship Statement

All authors have reviewed and approved the final manuscript. All listed authors meet authorship criteria and that no other individuals meeting the criteria were omitted. JEM acted as the guarantor of this paper.

PYN: conceptualisation, data curation, formal analysis, investigation, methodology, writing – original draft preparation

EK: investigation, validation, writing – review and editing

MJP, SLT: investigation, validation, supervision, writing – review and editing

JEM: conceptualisation, methodology, validation, formal analysis, supervision, writing – review and editing

