## Supplementary Files for "Selective reporting of outcomes and results in interrupted time series studies of health interventions: a methodological study"

**S1. Deviations from protocol**

| Original plan | Modified plan | Reason for modification |
|---|---|---|
| We planned to reproduce all meta-analyses in the sample. | We did not attempt reproduction for meta-analyses generated using Review Manager software, given the forest plot shows all model details and summary statistics or effect estimates and measures of precision. We assumed that such meta-analyses are inherently reproducible. | The forest plots generated by Review Manager often display most of the statistical details necessary to reproduce the meta-analysis accurately, and present the necessary study-level data. From previously working with Review Manager, we anticipate that reproducing such meta-analyses would always yield the same result as the original. |
| We planned to present descriptive statistics for all systematic reviews in the sample. | We presented descriptive statistics for the whole sample of systematic reviews, as well as for the subsample of systematic reviews in which meta-analyses were generated using software other than Review Manager (RevMan). | We were interested in determining whether our results were driven by meta-analyses generated using RevMan or via other software. |
| We planned to reproduce the meta-analysis using the *metafor* package in R software if no analytic code was shared or if we were unable to | In these scenarios, we reproduced the meta-analysis using the *meta* package in R. | Based on the 20 reviews that specified the R package used in our sample, the *meta* package was used more often (n=13) than the *metafor* package (n=10). We hypothesized |

| Original plan | Modified plan | Reason for modification |
| --- | --- | --- |
| access the software package used by the original systematic reviewers. | | that using the *meta* package would increase the likelihood of consistent use of the package between the original and reproduced meta-analysis, which would reduce any differences in results that could occur due to variations in statistical algorithm between packages. |
| We planned to consider a meta-analysis as reproducible regardless of whether summary statistics or effect estimates were used in reproducing the meta-analysis. | We classified whether the reproduction was achievable using the summary statistics or just the effect estimates for individual studies and presented that information on the Bland-Altman plots. | Although meta-analysis using only effect estimates was possible, the reproduced meta-analysis would be restricted to the generic inverse variance method only, which may deviate from the methods of the original meta-analysis (e.g. if the authors had used summary statistics to generate meta-analyses, but only presented effect estimates and measures of precision on the forest plot). Therefore, presenting this information on the Bland-Altman plots allows us to highlight the challenges in obtaining and using summary statistics in reproducibility studies. |
| We planned to classify index meta-analyses as 'Results fully reproducible' when there was no difference [with allowance for trivial discrepancies such as those due to computational algorithms] observed between the | We classified index meta-analyses as 'Results fully reproducible' when there was less than a 10% difference observed between the original and recalculated summary effect | To standardise our judgement of whether a difference was "trivial", we adopted a 10% threshold. Also we did not compare inferences about heterogeneity (e.g. I2 statistic) between the original and reproduced meta-analysis, given such inferences are heavily influenced by the |

| Original plan | Modified plan | Reason for modification |
|---|---|---|
| original and recalculated meta-analytic effect estimate, its 95% confidence interval and inferences about heterogeneity reported in the original review. | estimate and its 95% confidence interval's width reported in the original review. | heterogeneity estimator used, which often needed to be inferred |
| We planned for investigators involved in the reanalyses to specify whether they believe the observed difference between the original and recalculated summary estimate and its precision was meaningful, that is, would lead to a change in the interpretation of the results (classified as 'difference meaningful' or 'difference not meaningful'). | We classified a difference as 'meaningful' as any of the following: the original and reproduced summary effect estimates had the opposite direction of effect (i.e. above versus below the null value); or the original 95% CI included the null while the reproduced 95% CI did not include the null, or vice versa. | To standardise our judgement of whether the difference was "meaningful", we defined scenarios that would lead to such judgements. |

**S2. Template for correspondence with systematic reviewers**

**2.1. Template of invitation email**

**Subject: Data request for REPRISE study evaluating reproducibility of meta-analyses**

Dear [name],

My name is Phoebe Nguyen, and I am a Research Officer based at Monash University (Melbourne, Australia). I'm currently undertaking research on the reproducibility of meta-analyses, which explores how often we get the same meta-analysis results when we repeat the analysis on the same dataset. This research is funded through an Australian Research Council Discovery Early Career Researcher Award (DE200101618) and has been approved by the Monash University Human Research Ethics Committee (approval number: 30538).

As part of this research, we have assembled a random sample of systematic reviews with meta-analysis of the effects of a health, social, behavioural or educational intervention. We are contacting the authors of the included systematic reviews to ask if they would be willing to share data from one meta-analysis in their review (please see below for specific details). We plan to evaluate the reproducibility of these meta-analyses, by reanalysing them using the same methods as used by the original review authors and checking how consistent the findings are. We will seek input from the original review authors if we observe any discrepancies upon reanalysis, and report in aggregate the frequency of discrepancies between the original and re-analysed meta-analysis results. A protocol for our study has been published (doi: 10.1186/s13643-021-01670-0).

Following our reproducibility checks, we plan to create a publicly available repository of meta-analysis datasets that can be used by others for teaching purposes and illustration/testing of new meta-analytic methods.

<u>What we are seeking from you</u>

Your systematic review, *[title of article]*, published in *[journal]* in 2020, has been included in our random sample and provides data that we would like to include in our study. Specifically, we are seeking a file containing the data presented in **[figure/table reference]** for the outcome **[name of outcome],** and, if applicable, **the analytic code** used to generate the meta-analysis.

If you are willing to share your data file, please email it in a file format convenient for you (e.g. xlsx, csv, dta). If you'd prefer, we are happy to provide you with a link to a private Google Drive Folder which you can upload the file to. The variables that we require in the file are summary statistics for each group for individual studies (e.g. number of events, sample size), or effect estimates (e.g. risk ratio) and measures of precision (e.g. 95% confidence interval or standard error) for individual studies, or all of the above if readily available.

An example of the data file format for a meta-analysis of a binary outcome follows:

| Study ID | Intervention Events | Intervention Total | Control Events | Control Total | Risk ratio | 95%CI lower | 95%CI upper |
|---|---|---|---|---|---|---|---|
| 1 | 37 | 141 | 17 | 52 | 0.80 | 0.50 | 1.29 |
| 2 | 41 | 137 | 16 | 49 | 0.92 | 0.57 | 1.48 |
| 3 | 7 | 35 | 5 | 29 | 1.16 | 0.41 | 3.27 |
| … | … | | | | | | |

If you wrote code/scripts to analyse the data and are willing to share it, please email the code in a file format convenient to you (e.g. txt, R, do).

Consent

To assist in your decision to participate, we provide the Explanatory Statement attached. We have also attached a Consent Form, on which you may indicate your consent to the use of your provided dataset in our re-analysis and for sharing it via the Open Science Framework online repository.

We are grateful for your time in contributing to this project. Please do not hesitate to contact me if you have any questions.

With best wishes,

Phoebe Nguyen

REPRISE project manager, Monash University, Australia

On behalf of the REPRISE project investigator team: Matthew Page (Monash University, Australia), Joanne McKenzie (Monash University, Australia), David Moher (Ottawa Hospital Research Institute, Canada), Fiona Fidler (University of Melbourne, Australia), Julian Higgins (University of Bristol, UK), Sue Brennan (Monash University, Australia), Neal Haddaway (Leibniz-Centre for Agricultural Landscape Research, Germany), Daniel Hamilton (University of Melbourne, Australia), Raju Kanukula (Monash University, Australia), Sathya Karunananthan (McGill University, Canada), Lara Maxwell (University of Ottawa, Canada), Steve McDonald (Monash University, Australia), Shinichi Nakagawa (University of New South Wales, Australia), David Nunan (Oxford University, UK), Peter Tugwell (University of Ottawa, Canada), Vivian Welch (Bruyère Research Institute, Canada)

#### 2.2. Explanatory Statement

### EXPLANATORY STATEMENT

**Project:** Evaluation of the reproducibility of meta-analyses (the REPRISE project)

**Chief Investigator:** Dr Matthew Page
School of Public Health and Preventive Medicine, Monash University

You are invited to take part in this study. Please read this Explanatory Statement in full before deciding whether to participate in this research. If you would like further information regarding any aspect of this project, you are encouraged to contact the researchers via the phone number or email address listed above.

**What does the research involve?**

Reproducibility of results – that is, obtaining the same results when reanalysing the same dataset using the same computational methods – is considered by many to be an essential part of the scientific method. However, reproducibility is rarely discussed in relation to systematic reviews with meta-analysis, which focus on the totality of available evidence for a given question. To address this, we are undertaking a study to evaluate the reproducibility of meta-analyses, that is, the extent of variation in results when we independently re-run meta-analyses using the same computational steps and analytic code (if available) as used in a sample of original systematic reviews.

We are contacting you to ask if you are willing to share data (and if applicable, analytic code) for one meta-analysis in a systematic review you conducted. We plan to evaluate the reproducibility of the meta-analysis, by reanalysing it using the same methods as originally used by you and checking how consistent the findings are. Following our reproducibility checks, we plan to create a publicly available repository of meta-analysis datasets that can be used by others for teaching purposes and illustration/testing of new meta-analytic methods. This repository would include, for each study included in the meta-analysis, a study identifier, study citation, outcome description, summary statistics for each group, and effect estimates and measures of precision.

**Why were you chosen for this research?**

You have been invited because you were a corresponding author of a systematic review published in 2020.

**Who is funding this research?**

The project is funded by an Australian Research Council Discovery Early Career Researcher Award (DE200101618).

**Consenting to participate in the project and withdrawing from the research**

If you are willing to share your data for use in this project, please complete the consent form attached to the invitation email. Providing data for this study is voluntary, as is consenting for the data to be shared via an online data repository. If you consent to the use of your data in any aspect of the project, you may choose to withdraw consent at any stage prior to analysis or creation of the repository.

**Possible benefits and risks to participants**

The main output of this project will be a greater understanding of the extent to which results of meta-analyses are reproducible, which we anticipate will inform future methodological guidance on how best to report methods and results. For the community of systematic review methodologists and statisticians, the compiled open access repository of meta-analysis datasets will allow future methodological research to be undertaken. For the wider community, we hope that the knowledge gained will ultimately lead to higher quality systematic reviews with meta-analysis, thus facilitating evidence-based decision-making.

The risks to participants involved in this research are small. It is possible that we may identify potential errors in the data you share, in which case we will contact you to clarify them. We will report in aggregate, not individually, the frequency of discrepancies between the original and re-analysed meta-analysis results.

**Confidentiality**

In our comparison of the original meta-analysis results with the reanalysed meta-analysis results, results from your dataset will not be identifiable. The data we share via the publicly available repository will be identified as coming from your systematic review through inclusion of the review citation in the repository.

**Storage of data**

Data will be collected via email or via provision of a link to a private Google Drive Folder, unique to you. Access to the Folder will be restricted to yourself and the chief investigator (as listed above). At the conclusion of the project, data will be transferred to a Monash University secure server, where they will be stored for five years in accordance with Monash University regulations. In addition, a CSV-formatted spreadsheet containing aggregate data for all meta-analyses reanalysed by the study investigators will be deposited and stored indefinitely on the Open Science Framework, which is supported through an in-perpetuity trust.

**Use of data for other purposes**

With your consent, your dataset may be used in future research projects.

**Results**

Results of the study will be submitted for publication in an open-access, peer-reviewed journal and presented at conferences.

**Complaints**

Should you have any concerns or complaints about the conduct of the project, you are welcome to contact the Executive Officer, Monash University Human Research Ethics Committee (MUHREC).

Thank you,

Dr Matthew Page, PhD

Project lead and Senior Research Fellow, Monash University, Australia

##### 2.3 Consent form

#### CONSENT FORM

Project: Evaluation of the reproducibility of meta-analyses (the REPRISE project)

Chief Investigator: Dr Matthew Page, School of Public Health and Preventive Medicine, Monash University.

I have been asked to take part in the Monash University research project specified above. I have read and understood the Explanatory Statement and I hereby consent to participate in this project.

| I consent to the following: | Yes |
|---|---|
| The data that I provide may be used in the reproducibility evaluation | ☐ |
| The data that I provide may be deposited in the Open Science Framework online data repository | ☐ |
| The data that I provide may be used in future research projects (with appropriate ethics approval) | ☐ |

Name of participant:

Participant signature:

Date:

This study has been approved by the Monash University Human Research Ethics Committee.

The Approval number is: 30538

If you have any complaints or reservations about the ethical conduct of this research, you may contact the Executive Officer, Monash University Human Research Ethics Committee.

**S3. Template code for replication of meta-analysis**

Highlighted section is customised to the statistical specifications of each reproduction.

| | |
|---|---|
| Stata<br>*meta* command | /*Meta-analysis of continuous data, with summary statistics*/<br>version xx // replace xx with version number<br>import delimited "C:\Datafile.csv", clear // replace with link to data file<br>meta esize en emean esd cn cmean csd, esize(mdiff)studylabel(id) nometashow<br>meta summarize, cformat(%9.4f) pformat(%5.4f) sformat(%8.4f)<br><br>/*Meta-analysis of binary data, with summary statistics*/<br>version xx<br>import delimited "C:\Datafile.csv", clear<br>meta esize ecase enoncase ccase cnoncase, esize(lnoratio) fixed(iv) studylabel(id) nometashow<br>meta summarize, eform cformat(%9.4f) pformat(%5.4f) sformat(%8.4f)<br><br>/*Meta-analysis of continuous data, without summary statistics*/<br>version xx<br>import delimited "C:\Datafile.csv", clear<br>meta set ee cill ciul, random(dlaird) studylabel(id) cformat(%9.4f) pformat(%5.4f) sformat(%8.4f)<br>meta summarize, eform cformat(%9.4f) pformat(%5.4f) sformat(%8.4f)<br><br>/*Meta-analysis of binary data, without summary statistics*/<br>version xx<br>import delimited "C:\Datafile.csv", clear<br>meta set lnee lncill lnciul, random(dlaird) studylabel(id) civartolerance(1e-5)<br>meta summarize, eform subgroup(subgroup) cformat(%9.4f) pformat(%5.4f) sformat(%8.4f) |
| Stata<br>*metan* command | /*Meta-analysis of continuous data, with summary statistics*/<br>version xx |

| | |
|---|---|
| | import delimited "C:\Datafile.csv", clear<br>metan en emean esd cn cmean csd, hedges random(dlaird) lcols(id) nograph<br>return list<br>matrix list r(ovstats)<br><br>/*Meta-analysis of binary data, with summary statistics*/<br>version xx<br>import delimited "C:\Datafile.csv", clear<br>metan ecase enoncase ccase cnoncase, or random(dlaird) lcols(id) nograph<br>return list<br>matrix list r(ovstats)<br><br>/*Meta-analysis of continuous data, without summary statistics*/<br>version xx<br>import delimited "C:\Datafile.csv", clear<br>metan lnee lncill lnciul, random(dlaird) lcols(id) eform nograph<br>return list<br>matrix list r(ovstats)<br><br>/*Meta-analysis of binary data, without summary statistics*/<br>version xx<br>import delimited "C:\Datafile.csv", clear<br>metan ee cill ciul, random(dlaird) lcols(id) eform nograph<br>return list<br>matrix list r(ovstats) |
| R<br>*meta* package | library(meta)<br>library(grid) |

| | |
|---|---|
|  | library(readr)<br><br>path <- "C:\\Datafile.csv" #replace with link to data file<br>dat<-read_csv(path)<br><br>## Meta-analysis of continuous data, with summary statistics<br>metadat <- metacont(en,emean,esd,cn,cmean,csd,<br> data=dat,<br> fixed=FALSE,random=TRUE,<br> method.tau='DL',<br> sm="SMD",method.smd="Hedge",<br> label.e="Intervention",label.c="Control",<br> overall=TRUE,overall.hetstat=TRUE,<br> subgroup=subgroup,<br> studlab=id)<br>summary(metadat)<br><br>## Meta-analysis of binary data, with summary statistics<br>metadat <- metabin(ecase,en,ccase,cn,<br> data=dat,<br> fixed=FALSE,random=TRUE,<br> method="Inverse",<br> method.tau='DL',<br> sm="OR",<br> label.e="Intervention",label.c="Control",<br> overall=TRUE,overall.hetstat=TRUE,<br> subgroup=subgroup,<br> studlab=id) |

```
summary(metadat)

##Meta-analysis of continuous data, without summary statistics
metadat <- metagen(data=dat,
                TE=smd,
                seTE=se,
                lower=cill,upper=ciul,
                studlab=id,
                subgroup=subgroup,
                sm="SMD",
                fixed=FALSE,random=TRUE,
                method.tau='DL',
                label.left="Intervention",label.right="",
                overall=TRUE,overall.hetstat=TRUE)
summary(metadat)

##Meta-analysis of binary data, without summary statistics
metadat <- metagen(data=dat,
                TE=lnor,
                seTE=se,
                lower=lncill,upper=lnciul,
                studlab=id,
                subgroup=subgroup,
                sm="OR",
                fixed=FALSE,random=TRUE,
                method.tau='DL',
                label.left="Intervention",label.right="",
                overall=TRUE,overall.hetstat=TRUE)
```

| | |
|---|---|
| | summary(metadat) |
| R<br>*metafor* package | library(metafor)<br>library(grid)<br>library(readr)<br>library(dplyr)<br><br>path <- "C:\\Datafile.csv"<br>dat<-read_csv(path)<br>dat<-dat %>% mutate(sei=(ciul-cill)/3.92)<br><br>#Random-effects model<br>ma <- rma.uni(yi=smd,sei=sei,data=dat,method=’DL’)<br>summary(ma)<br><br>#Fixed-effect model<br>ma <- rma.uni(yi=smd,sei=sei,data=dat,method=’FE’)<br>summary(ma)<br><br>#Subgroup meta-analysis<br>ma <- rma.uni(yi=smd,sei=sei,data=dat,method=’REML’)<br>ma1 <- rma.uni(yi=smd,sei=sei,data=dat,method=’REML’,subset=(subgroup==”Subgroup 1”))<br>ma2 <- rma.uni(yi=smd,sei=sei,data=dat,method=’REML’,subset=(subgroup==”Subgroup 2”))<br>summary(ma)<br>summary(ma1)<br>summary(ma2) |

#### S4. Additional details on how reproduction of meta-analyses were performed

During the period of reproduction (February to October 2022), the versions of the software used at the time of reproduction were as follows: R v4.1.3 to v4.2.2 (*meta* v5.2-0 to v6.0-0; *metafor* v3.4-0 to v3.8-1), Stata (set to the version used in the original study, or v15.0 if not specified), and Comprehensive Meta-Analysis (CMA) v4.

##### 4.1 Transformation of data prior to replication

For meta-analyses of standardised mean differences (SMD):

- Difference in SMD (ΔSMD) = reproduced SMD – original SMD
- Standard error (SE) of ΔSMD = (upper limit of original 95% CI – lower limit of original 95% CI) ÷ 3.92
- Difference in width of 95% confidence interval (ΔCI) = reproduced CI width – original CI width

For meta-analyses of odds ratios (OR):

- Ratio of OR (rOR) = ln(reproduced OR) – ln(original OR)
- SE of rOR = [ln(upper limit of original 95% CI) – ln(lower limit of original 95% CI)] ÷ 3.92
- Difference in width of 95% confidence interval (ΔCI) = reproduced CI width – original CI width

For meta-analyses of risk ratios (RR):

- Ratio of RR (rRR) = ln(reproduced RR) – ln(original RR)
- SE of rRR = [ln(upper limit of original 95% CI) – ln(lower limit of original 95% CI)] ÷ 3.92
- Difference in width of 95% confidence interval (ΔCI) = reproduced CI width – original CI width

For meta-analyses of hazard ratios (RR):

- Ratio of HR (rHR) = ln(reproduced HR) – ln(original HR)
- SE of rHR = [ln(upper limit of original 95% CI) – ln(lower limit of original 95% CI)] ÷ 3.92
- Difference in width of 95% confidence interval (ΔCI) = reproduced CI width – original CI width

For meta-analyses of OR, RR and HR, if any of the original CI limits were reported as zero, we used the RevMan v5.4 calculator to generate that value, ensuring that the 95% CI remained symmetrical around the effect estimate. For example, if the lower limit of the 95% CI was reported as 0, the mean and the upper limit of the 95% CI was inputted into RevMan calculator to generate the value of the lower limit.

All reproduced results were rounded to the same decimal points as the original results, before their discrepancies were calculated. Limits of confidence intervals were rounded separately and the rounded values were used to calculate the confidence interval width, which was then compared with the original confidence interval width.

#### 4.2 Algorithm for selection of statistical methods

[redacted]

[redacted]

[redacted]

Note: For studies conducted in Stata without specifying the command used, we used the *metan* command since the *meta* command was not available for Stata version 15 and earlier.

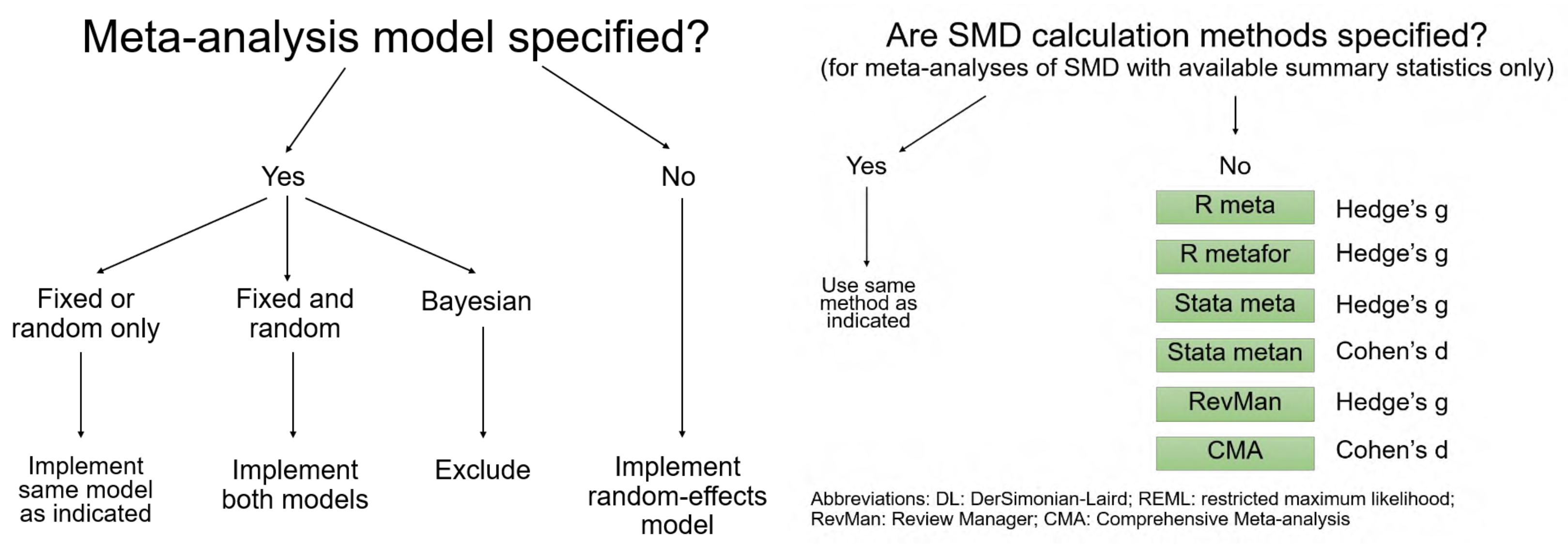
Meta-analysis model specified?
Yes
No
Fixed or random only
Fixed and random
Bayesian
Implement same model as indicated
Implement both models
Exclude
Implement random-effects model
Are SMD calculation methods specified?
(for meta-analyses of SMD with available summary statistics only)
Yes
No
Use same method as indicated
R meta
Hedge's g
R metafor
Hedge's g
Stata meta
Hedge's g
Stata metan
Cohen's d
RevMan
Hedge's g
CMA
Cohen's d
Abbreviations: DL: DerSimonian-Laird; REML: restricted maximum likelihood; RevMan: Review Manager; CMA: Comprehensive Meta-analysis

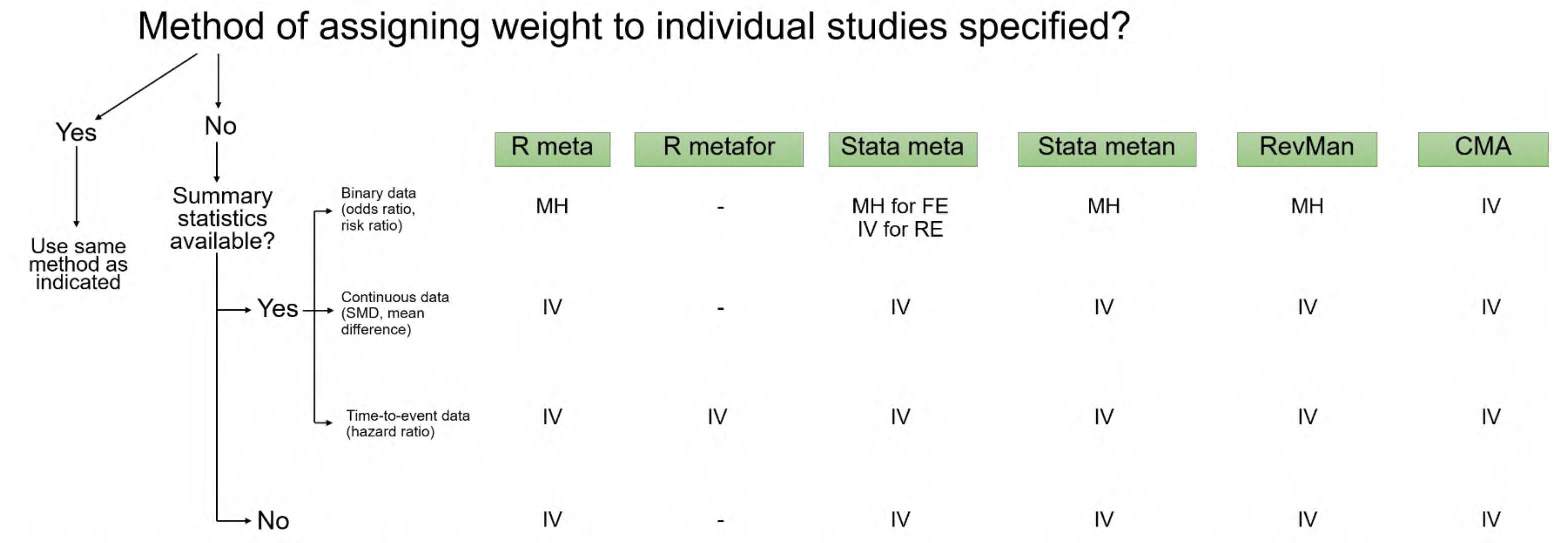


Abbreviations: IV: inverse variance; MH: Mantel-Haenszel; RevMan: Review Manager; CMA: Comprehensive Meta-analysis; FE/RE: fixed-effect/random-effects

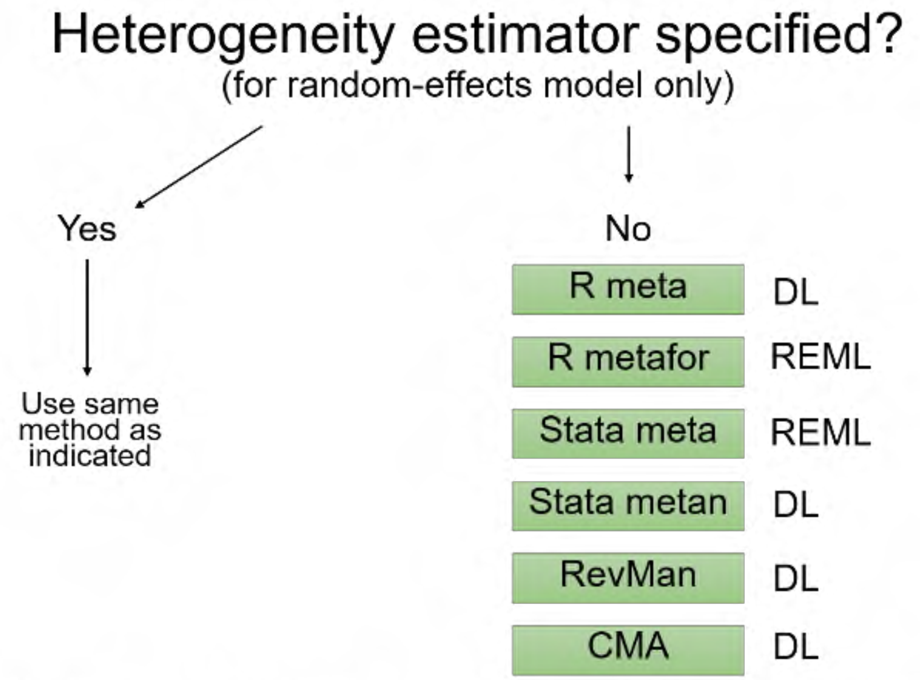


Abbreviations: DL: DerSimonian-Laird; REML: restricted maximum likelihood; RevMan: Review Manager; CMA: Comprehensive Meta-analysis

For R *meta* package, DerSimonian-Laird was the default estimator for all versions earlier than 5.0-0 of the package (i.e. before 11/10/2021).

#### S5. Flow diagram of study identification, screening and inclusion

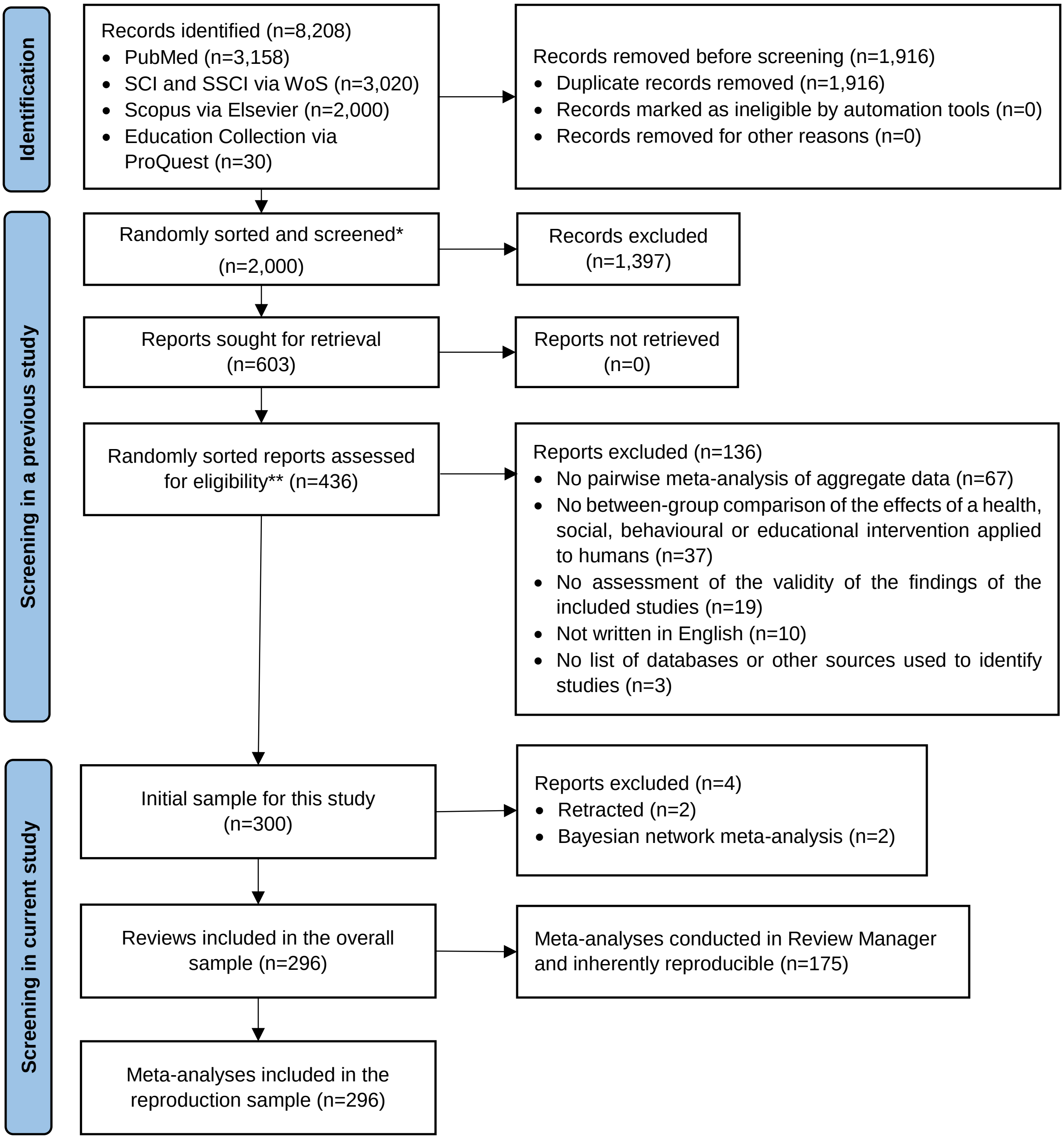


**There were 6,292 unique records after duplicates were removed, but we only needed to screen the first 2000 randomly sorted records to reach our target sample size.*

*The full process of study identification and screening to obtain the initial sample (n=300) was described in full details in this paper: Nguyen PY, Kanukula R, McKenzie JE, et al. Changing patterns in reporting and sharing of review data in systematic reviews with meta-analysis of the effects of interventions: cross sectional meta-research study. bmj. 2022 Nov 22;379.*

#### S6. Frequency of reporting of statistical details and meta-analysis results

| Item | Frequency (%) | |
|---|---|---|
| | All reviews | Reviews not using Review Manager |
| **Reporting of statistical details** | | |
| Software used for meta-analysis | | |
| Reported in article | 291/296 (98%) | 116/121 (96%) |
| Provided in author's correspondence | 1/296 (<0.5%) | 1/121 (1%) |
| Not determinable | 4/296 (1%) | 4/121 (3%) |
| Software version | 262/296 (89%) | 100/121 (83%) |
| Package or command used for meta-analysis (among reviews using R or Stata) | | |
| Reported in article | 28/85 (33%) | 28/85 (33%) |
| Provided in author's correspondence | 7/85 (8%) | 7/85 (8%) |
| Not determinable | 50/85 (59%) | 50/85 (59%) |
| Method of calculating SMD (among reviews with meta-analyses of SMD) | | |
| Reported in article | 17/62 (27%) | 14/31 (45%) |
| Provided in author's correspondence | 31/62 (50%) | 3/31 (10%) |
| Not determinable | 14/62 (23%) | 14/31 (45%) |
| Statistical model used for meta-analysis | | |
| Reported in article | 293/296 (99%) | 118/121 (98%) |
| Provided in author's correspondence | 0/296 (0%) | 0/121 (0%) |
| Not determinable | 3/296 (1%) | 3/121 (2%) |
| Method of assigning weight to individual study | | |
| Reported in article | 208/296 (70%) | 33/121 (27%) |
| Provided in author's correspondence | 5/296 (2%) | 5/121 (4%) |
| Not determinable | 83/296 (28%) | 83/121 (69%) |

| Item | Frequency (%) | |
|---|---|---|
| | All reviews | Reviews not using Review Manager |
| Heterogeneity variance estimator (among reviews with random-effects model) | | |
| Reported in article | 47/222 (21%) | 35/103 (34%) |
| Provided in author's correspondence | 8/222 (4%) | 8/103 (8%) |
| Not determinable | 167/222 (75%) | 60/103 (58%) |
| Method of continuity correction (among reviews with zero-cell counts) | 5/36 (14%) | 2/9 (22%) |
| **Reporting of meta-analysis results** | | |
| Summary effect estimate | 296/296 (100%) | 121/121 (100%) |
| 95% confidence interval limits | 296/296 (100%) | 121/121 (100%) |
| P-value of the summary effect | 245/296 (83%) | 71/121 (59%) |
| As exact values | 171/296 (58%) | 44/121 (36%) |
| As a range (e.g. p<0.01) | 74/296 (25%) | 27/121 (22%) |
| Not reported | 51/296 (17%) | 50/121 (41%) |
| $I^2$ statistic | 284/296 (96%) | 109/121 (90%) |
| $Tau^2$ statistic | 148/222 (67%) | 30/103 (29%) |
| $Chi^2$ statistic of heterogeneity | 208/296 (70%) | 36/121 (30%) |

**S7. Sensitivity analysis | Using a 5% threshold to classify reproducibility instead of 10%**

| **No. meta-analyses** | **Using 5% threshold** | **Using 10% threshold** |
|---|---:|---:|
| Fully reproducible | 93 (77%) | 104 (86%) |
| Not fully reproducible | 18 (15%) | 7 (6%) |
| Results not able to be reproduced | 10 (8%) | 10 (8%) |
| **Total** | **121 (100%)** | **121 (100%)** |

**S8. Banksia plots comparing the scaled reproduced meta-analysis summary effect estimates (and 95% confidence intervals) against the original.**

**(A) Standardised mean difference**

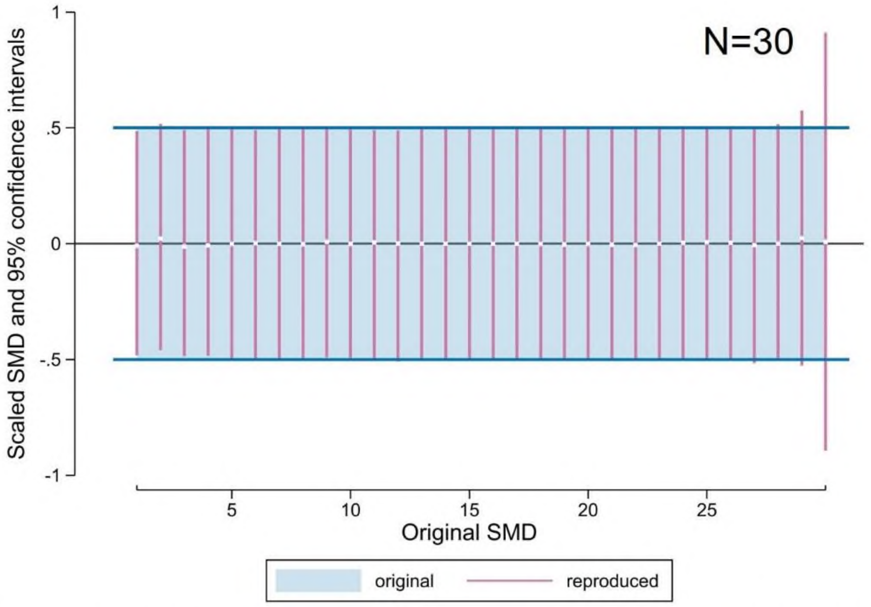


*Plot of reproduced meta-analysis summary estimates compared to the original estimates (the reference) in 30 meta-analyses of standardised mean differences (SMD). White squares represent the difference between the reproduced and the original meta-analysis summary estimates. Purple vertical lines represent the confidence intervals (CIs) of reproduced estimates on the scale of the original estimates (where the original estimate is set to 0 and its CI to -0.5; 0.5). CI lines are sorted by width, narrowest to the left. The horizontal black line indicates a value of scaled difference equal to zero (null value).*

**(B) Mean difference**

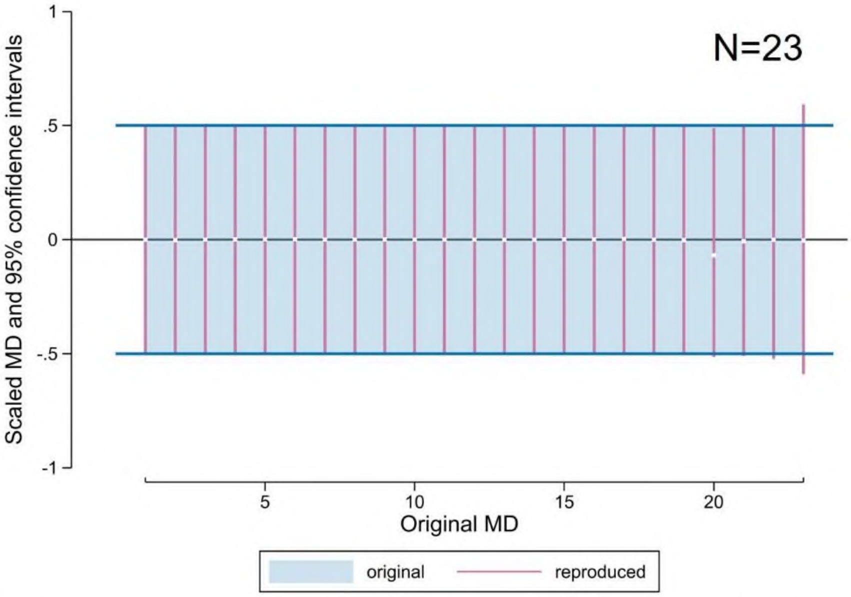


*Plot of reproduced meta-analysis summary estimates compared to the original estimates (the reference) in 23 meta-analyses of mean differences (MD). White squares represent the difference between the reproduced and the original meta-analysis summary estimates. Purple vertical lines represent the confidence intervals (CIs) of reproduced estimates on the scale of the original estimates (where the original estimate is set to 0 and its CI to -0.5; 0.5). CI lines are sorted by width, narrowest to the left. The horizontal black line indicates a value of scaled difference equal to zero (null value).*

**(C) Hazard ratio**

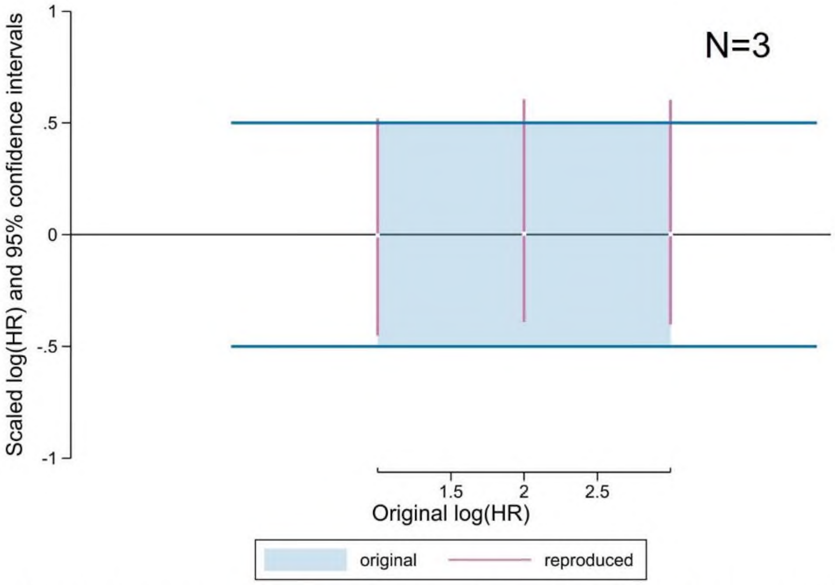


*Plot of reproduced meta-analysis summary estimates compared to the original estimates (the reference) in 3 meta-analyses of hazard ratios (HR). White squares represent the difference between the reproduced and the original meta-analysis summary estimates. Purple vertical lines represent the confidence intervals (CIs) of reproduced estimates on the scale of the original estimates (where the original estimate is set to 0 and its CI to -0.5; 0.5). CI lines are sorted by width, narrowest to the left. The horizontal black line indicates a value of scaled difference equal to zero (null value). The summary effect estimates and the CI are presented on a log scale.*

**(D) Odds ratio**

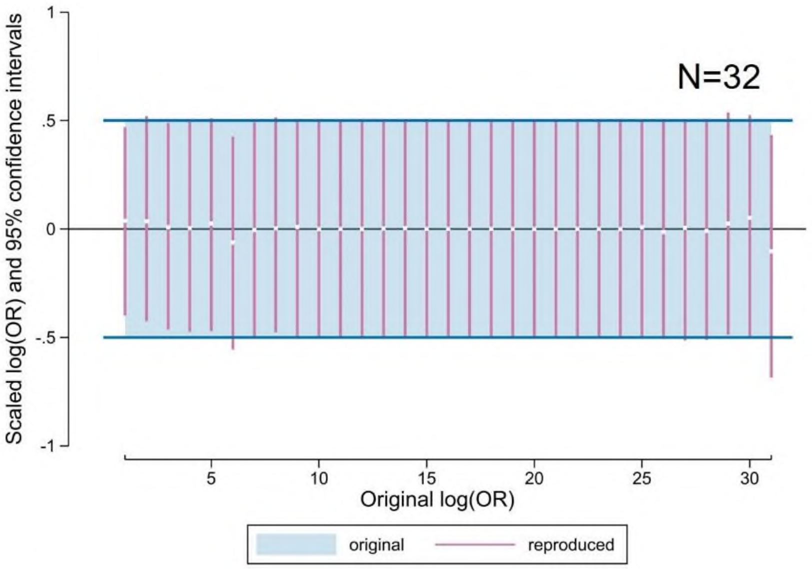


*Plot of reproduced meta-analysis summary estimates compared to the original estimates (the reference) in 31 meta-analyses of odds ratios (OR). White squares represent the difference between the reproduced and the original meta-analysis summary estimates. Purple vertical lines represent the confidence intervals (CIs) of reproduced estimates on the scale of the original estimates (where the original estimate is set to 0 and its CI to -0.5; 0.5). CI lines are sorted by width, narrowest to the left. The horizontal black line indicates a value of scaled difference equal to zero (null value). The summary effect estimates and the CI are presented on a log scale.*

**(E) Risk ratio**

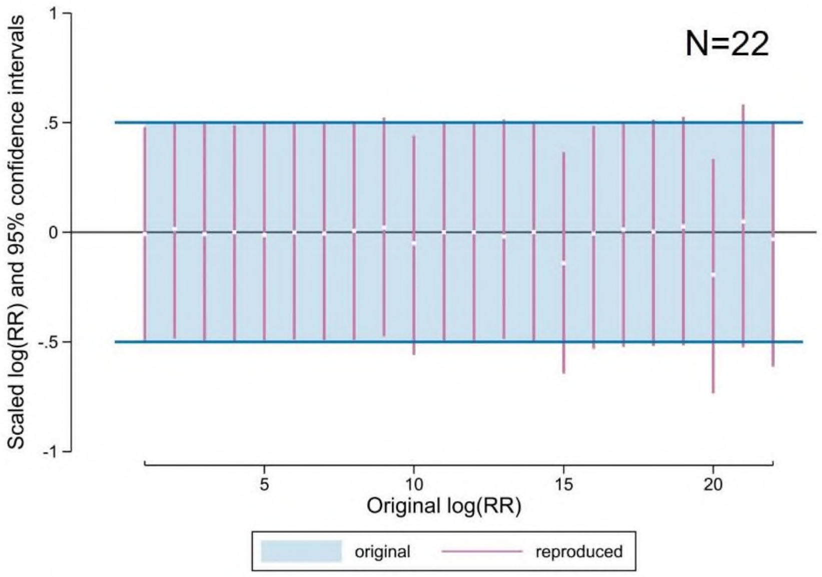


*Plot of reproduced meta-analysis summary estimates compared to the original estimates (the reference) in 22 meta-analyses of risk ratios (RR). White squares represent the difference between the reproduced and the original meta-analysis summary estimates. Purple vertical lines represent the confidence intervals (CIs) of reproduced estimates on the scale of the original estimates (where the original estimate is set to 0 and its CI to -0.5; 0.5). CI lines are sorted by width, narrowest to the left. The horizontal black line indicates a value of scaled difference equal to zero (null value). The summary effect estimates and the CI are presented on a log scale.*

**About the Banksia plot**

Each original summary effect estimate was treated as the reference value and centred on zero, and its confidence interval was scaled to span a range of one (from -0.5 to 0.5). Its corresponding reproduced summary effect estimate and confidence interval was then adjusted by the same amount. This allows easy identification of whether the scaled differences and their confidence intervals are similar or different between the original and reproduced summary estimates. Plots were generated separately for meta-analyses of SMD, log(RR), log(OR) and log(HR).

Information on how to construct and interpret the Banksia plot can be found in the following article:

*Turner SL, Karahalios A, Korevaar E, McKenzie JE. The Banksia plot: a method for visually comparing point estimates and confidence intervals across datasets. J Clin Epidemiol. 2025 Jan;177:111591.*

**S9. Bland-Altman plots comparing the reproduced and original meta-analysis summary effect estimates | Overall sample**

##### (A) Meta-analysis of standardised mean differences

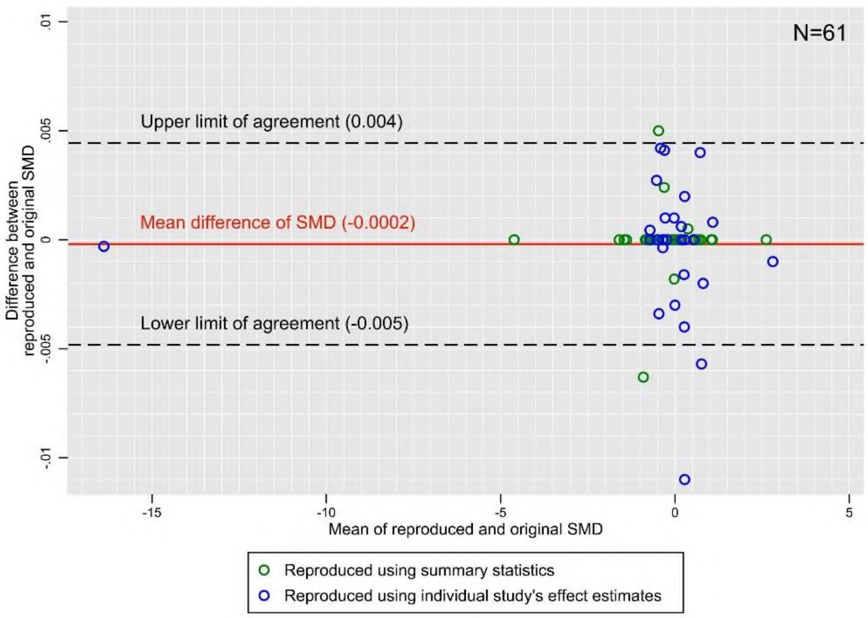


##### (B) Meta-analysis of hazard ratios

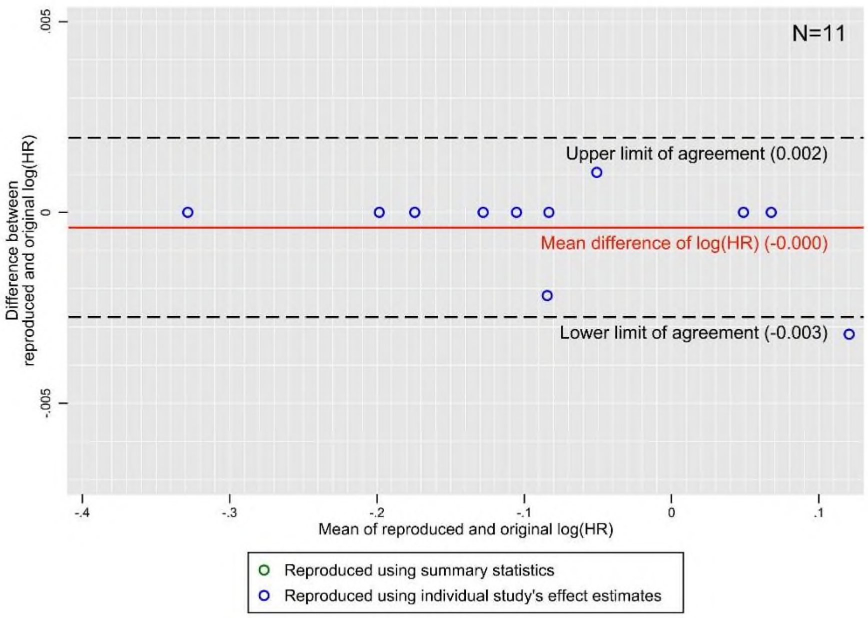

#### (C) Meta-analysis of odds ratios

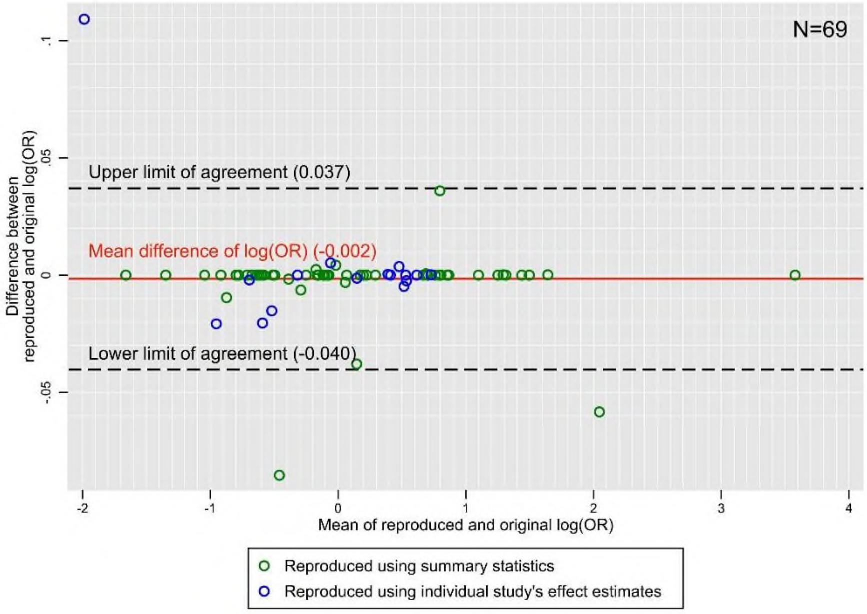


#### (D) Meta-analysis of risk ratios

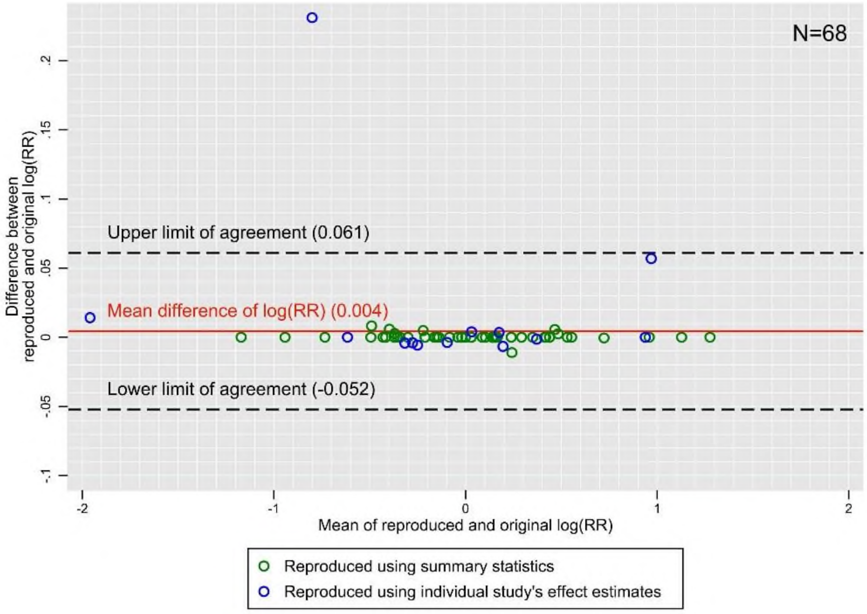
